# Enhanced screening and bacterial sexually-transmitted infection diagnoses after HIV pre-exposure prophylaxis initiation

**DOI:** 10.64898/2025.12.19.25342713

**Authors:** Anna M. Parker, Samuel M. Jenness, Benjamin J. Singer, Jennifer J. Chang, Katia J. Bruxvoort, Joseph A. Lewnard

**Author notes:** Address for correspondence: Joseph Lewnard, 2121 Berkeley Way, Room 5410, Berkeley, California 94122 United States. These authors jointly supervised and contributed equally to the work (Katia J. Bruxvoort, Joseph A. Lewnard). **Conflicts of interest:** JAL discloses receipt of grant funding from Gilead Sciences, Inc. The remaining authors declare no conflicts of interest. **Data availability:** Raw data from the MarketScan insurance claims databases are available for licensed users. A user license could be obtained by following the instructions at https://marketscan.truvenhealth.com/marketscanportal/.

## Abstract

**Background:** Recipients of HIV pre-exposure prophylaxis (PrEP) experience higher rates of gonorrhea and chlamydia diagnoses than non-recipients. However, it is unclear if these observations reflect a causal relationship between PrEP initiation and acquisition of sexually transmitted infections (i.e., behavioral “risk compensation”), or alternatively a diagnostic bias related to PrEP recipients screening more frequently than non-recipients.

**Methods:** We conducted a self-controlled case series study comparing rates of gonorrhea and chlamydia diagnoses after versus before PrEP initiation among commercially-insured US males in the Merative MarketScan® Research Databases (2016–2019). We compared matched incidence rate ratio (IRR) estimates after versus before PrEP initiation to model-based expectations under a null hypothesis of no change in infection risk. We derived null expectations via a mathematical model reflecting changes in screening after PrEP initiation, estimating parameters under a Bayesian framework.

**Results:** Individuals experienced increased rates of gonorrhea (IRR=3.47 [95% confidence interval: 2.57-4.72]) and chlamydia (IRR=3.59 [2.50-5.27]) diagnoses after PrEP initiation, with IRR estimates differing by anatomical site (IRR for either infection=1.23 [0.71-2.13] and 5.98 [3.63-10.29] for urogenital and extragenital diagnoses, respectively). After accounting for increased rates of screening after PrEP initiation, rates of gonorrhea and chlamydia diagnoses only modestly exceeded expectations under the null hypothesis of no change in infection risk (8-14% greater-than-expected rates of gonorrhea and chlamydia diagnoses, respectively; one-sided *p*>0.1 for all tests). Overall, we estimated that 80-81% of observed increases in rates of gonorrhea and chlamydia diagnoses after PrEP initiation, respectively, were attributable to increases in asymptomatic screening.

**Conclusions:** Our findings suggest higher-frequency asymptomatic screening, rather than behavioral risk compensation, is the primary driver of increased rates of gonorrhea and chlamydia diagnoses after PrEP initiation.

## INTRODUCTION

Pre-exposure prophylaxis (PrEP) with tenofovir disoproxil fumarate/emtricitabine (TDF-FTC) or emtricitabine/tenofovir-alafenamide (F-TAF) prevents HIV infection among men who have sex with men (MSM),^1–3^ who account for the vast majority of PrEP recipients in the United States (US).^4^ Among MSM, PrEP receipt is associated with greater frequency of condomless anal sex partnerships^5,6^ as well as positive psychosocial outcomes including reduced anxiety^7^ and increased sexual satisfaction and intimacy.^8^ However, reduced condom use and other forms of sexual behavior change after initiating PrEP may increase individuals’ risk of acquiring sexually-transmitted infections (STIs) besides HIV.

Whereas annual STI screening is recommended for most US MSM,^9^ those receiving PrEP are recommended to receive asymptomatic STI screening every 3-6 months.^10^ Three meta-analyses have identified elevated risk of STI diagnoses among MSM receiving PrEP,^11–13^ but causal implications of this observation are uncertain. Recipients of PrEP report greater numbers of sex partners, greater frequency of prior STI diagnoses, and greater STI testing frequency than PrEP non-recipients.^14–17^ These observations may reflect differences in sexual risk behaviors among individuals who initiate PrEP versus those who do not, or behavioral changes (“risk compensation”) resulting from individuals’ awareness of being at lower risk of HIV infection after initiating PrEP. Alternatively, increases in STI screening frequency after PrEP initiation may lead to increased rates of STI diagnoses absent any change in individuals’ risk of acquiring STIs. Modeling analyses have demonstrated that enhanced screening among PrEP recipients may offset increases in STI incidence expected to result from risk compensation.^18^ While several observational studies have sought to adjust for STI testing frequency when estimating changes in STI risk after PrEP adoption,^16,19,20^ standard adjustment frameworks risk introducing collider bias, as STI testing may be a consequence of both PrEP utilization and incident STIs (**Figure 1**).

**Figure 1:**
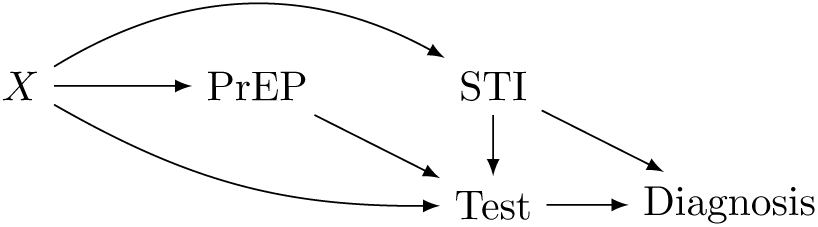
Directed acyclic graph. We illustrate a hypothesized causal pathway relating use of HIV pre-exposure prophylaxis (PrEP) to risk of sexually-transmitted infection (STI) diagnoses. While the causal relationship of interest is between PrEP use and risk of incident STIs, this effect can be measured only through STI diagnoses, as administrative data do not record all incident STIs. We consider that individuals’ frequency of STI testing is influenced by both PrEP use and incident STIs associated with symptoms. Rates of diagnoses are then a function of the frequency with which individuals acquire STIs and the frequency with which they are tested. We consider further that external risk factors (*X*), including calendar time but also potentially including time-invariant individual characteristics, influence individuals’ likelihood of initiating PrEP, their frequency of STI acquisition, and their frequency of testing. Standard frameworks adjusting for testing frequency may introduce collider bias, as testing may be a consequence of both PrEP use and incident STIs; administrative data do not provide a basis for distinguishing screening tests from those motivated by symptoms or suspicion of STI exposure. Our self-matched inference framework adjusts implicitly for time-invariant unobserved factors influencing individuals’ likelihood of PrEP initiation as well as their rates of STI acquisition and testing.

Understanding the contributions of risk compensation and screening practices to STI incidence rates among MSM receiving PrEP can inform the interpretation of trends in rates of STI diagnoses following PrEP implementation, as well as optimal testing recommendations for MSM receiving PrEP. We sought to distinguish the contribution of changes in infection risk and enhanced screening to changes in STI incidence after PrEP initiation. We aimed to mitigate potential sources of bias outlined above by quantifying within-individual changes in STI risk after PrEP initiation via a self-matched retrospective cohort study of commercially-insured US males initiating PrEP. We compared observed changes to expectations under the null hypothesis of no change in infection risk, which we parameterized via a Bayesian framework that accounted for changes STI screening frequencies.

## METHODS

### Design, study population, exposures, and outcomes

We undertook a self-matched, exposure-anchored (self-controlled case series) study among commercially-insured US males initiating PrEP between August 1, 2016 and October 31, 2019. The study population comprised members of insurance plans represented in the MerativeTM MarketScan® Research Databases, a US-wide multipayer collection of adjudicated claims data. Individuals eligible for inclusion in primary analyses were aged 18-64 years with male sex designated in member demographic characteristics, regardless of gender; who filled a ≥30-day supply of tenofovir disoproxil fumarate/emtricitabine (TDF-FTC) or emtricitabine/tenofovir-alafenamide (F-TAF) and who received a second ≥30-day TDF-FTC/F-TAF fill within the ensuing 365 days; who had ≥365 days of continuous membership before PrEP initiation (as defined below; **Table S1**) with no lapse in coverage ≥60 days; and who actively used their prescription plan (≥1 prescription fill within 365 days) before PrEP initiation. We defined the date of PrEP initiation as the first fill of a ≥30-day supply of TDF-FTC/F-TAF, or the first fill of any volume for patients who filled a ≥30-day supply within ≤90 days after a first fill of a smaller volume, as prescribers may start patients with a reduced volume to assess side effects. Following a previously-validated algorithm,^13^ we excluded individuals who may have received the same treatments for other reasons, including prior diagnosis of hepatitis B virus infection, HIV infection, or AIDS-defining illnesses (**Tables S2**-**S4**). Individuals receiving any needle stick diagnosis within 10 days before or after their first TDF-FTC/F-TAF fill (**Table S5**) were not considered to be initiating PrEP at this fill. Our analysis period preceded licensure of injectable cabotegravir or lenacapavir for PrEP; we therefore considered oral TDF-FTC/F-TAF fills only.

We compared incident STI diagnoses during risk periods comprising all eligible person-time after January 1^st^ 2016 through 30 days before PrEP initiation (control period), and beginning either 30 days after the date of PrEP initiation or at individuals’ second ≥30-day TDF-FTC/F-TAF fill, whichever occurred later (exposure period). We excluded observations from the end of the control period to the beginning of the exposure period as a washout interval because STIs diagnosed during this period could have been detected as a consequence of screening associated with PrEP initiation.^21^ Additionally, because recent STI diagnoses may be a factor in the decision to initiate PrEP,^22^ dates of PrEP initiation may cluster disproportionately within the period shortly following STI diagnoses, inflating apparent risk during the control period. To mitigate any resulting bias, we extended the washout interval through 6 months before PrEP initiation for individuals who received any STI diagnosis <6 months before initiating PrEP, or between their first and second PrEP fill. Sensitivity analyses described below modified washout and control period definitions. We defined the end of the exposure period as the earliest of the following events: 365 days after individuals’ most recent ≥30-day TDF-FTC/F-TAF fill; diagnosis with hepatitis B virus infection, HIV infection, or AIDS-defining illness; death; disenrollment (without re-enrollment within <60 days); or December 31, 2019.

We identified incident chlamydia and gonorrhea diagnoses via International Classification of Diseases, 10^th^ revision (ICD-10) codes (**Tables S6**-**S7**), defining new diagnoses as those occurring ≥30 days after any previous diagnosis involving the same pathogen. When ICD-10 codes specified an anatomical site (42% of diagnoses), we distinguished infections associated with any urogenital diagnosis from those associated with extragenital (rectal and pharyngeal) sites only, as urogenital infections are more likely to be associated with symptomatic illness in males.^23^ We also identified syphilis diagnoses via ICD-10 codes (**Table S8**). We considered syphilis diagnoses as incident infections if patients received penicillin within 10 days of diagnosis and had no prior syphilis diagnosis within ≥365 days. We present syphilis incidence for descriptive purposes only; diagnosed rates of syphilis infections and reinfections were prohibitively low for self-matched comparative analyses.

### Incidence rate ratios for STI diagnoses

We used conditional Poisson regression models^24^ to estimate self-matched incidence rate ratios (IRRs) comparing individuals’ risk of STI diagnoses during their exposure period versus their control period. To mitigate confounding driven by secular changes over time in STI prevalence and sexual behavior (including decreases in condom use among MSM not receiving PrEP^25^), we adjusted for time as a continuous variable. The self-matched inference framework was expected to mitigate confounding driven by time-invariant factors that may differ across individuals who initiate or do not initiate PrEP. We conducted statistical inference via bootstrap resampling.

We did several sensitivity analyses modifying eligibility criteria and person-time included in individuals’ control or exposure periods. These included analyses that reclassified the washout interval (from the date of the first TDF-FTC/F-TAF fill to the second fill) as unexposed person-time; analyses constraining the control period to 6 or 12 months before PrEP initiation, without washout for STI diagnoses during the control period; and analyses that did not require PrEP receipt to be associated with ≥2 fills. For the latter analyses, exposure periods began 30 days after individuals filled their first ≥30-day TDF-FTC/F-TAF supply.

Last, we conducted exploratory analyses aiming to understand other events associated with changes in individuals’ risk of STI diagnoses. To determine whether PrEP initiation was associated with persistent changes in STI risk after discontinuation, we defined a discontinuation period beginning 365 days after individuals’ last fill of a ≥30-day supply of TDF-FTC/F-TAF, and ending at any new fill; diagnosis with hepatitis B virus infection, HIV infection, or AIDS-defining illness; death; disenrollment (without re-enrollment within <60 days); or December 31, 2019. We estimated self-matched IRRs using conditional Poisson regression models adjusting for calendar time, consistent with primary analyses. To understand changes in STI incidence associated with other health events potentially triggering modifications in sexual risk behaviors or screening, we also assessed changes in risk of STI diagnoses after individuals were diagnosed with HIV infection. Eligible individuals were those aged 18-64 years; who had ≥365 days of continuous health plan membership before their first HIV diagnosis, with no lapse in coverage ≥60 days; and who were active users of their prescription plan (≥1 prescription fill within 365 days) before this diagnosis. We defined the control period as extending through an individuals’ first HIV diagnosis, and defined distinct risk periods within <6 months and ≥6 months after this diagnosis, considering that behavior may differ before and after establishment on antiretroviral treatment. We defined the end of each risk period as death, disenrollment (without re-enrollment within <60 days), or December 31, 2019.

### Distinguishing changes in rates of infection and screening

Whereas continued PrEP receipt may indicate frequent asymptomatic STI screening, administrative claims data do not distinguish STI screening tests from diagnostic testing for symptomatic illness. We leveraged the fact that extragenital infections are less likely to be symptomatic than urogenital infections to distinguish changes in rates of STI diagnoses attributable to changes in screening from changes in incident STI acquisition. We used a Bayesian framework to parameterize expected changes in rates of urogenital and extragenital STI diagnoses absent any change in infection risk with real-world data on testing frequency before and after PrEP initiation, as detailed below (**Model**). We conducted inference by comparing observed changes in rates of diagnoses to expectations under the null hypothesis of no change in infection, estimated using the fitted model (**Posterior**).

### Model

We formulated individuals’ rates of diagnoses, *D*, with pathogen *i* (corresponding to *N. gonorrhoeae* or *C. trachomatis*) by site (urogenital, 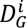, and extragenital, 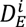) during the exposure period 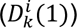 and control period 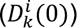. We considered the incidence rate of new urogenital infection diagnoses to represent the sum of two underlying rates: the rate of detection driven by clinical testing for new-onset symptomatic infections, and the rate driven by asymptomatic infection screening. We defined 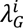 as the incidence rate of new urogenital infections, and *α^i^* as the proportion of urogenital infections causing symptoms and thus leading to clinical testing, yielding a rate of diagnoses of new-onset symptomatic urogenital infections equal to 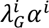. We defined 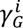 as the rate of spontaneous resolution of urogenital infections,^26–28^ and considered a pathogen-agnostic rate of screening for urogenital infections (σ*_G_*) as specimens are generally tested for both *N. gonorrhoeae* and *C. trachomatis*. Considering that asymptomatic infections could resolve spontaneously or due to treatment after being identified through screening, the expected prevalence of asymptomatic urogenital infections was 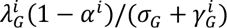, and the rate of detection of such infections was 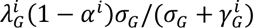. Thus,

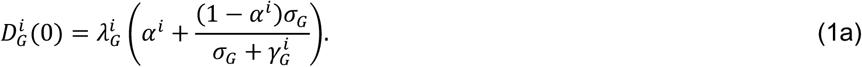

Assuming that extragenital infections are detected primarily due to screening, and defining 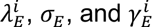 as the rates of extragenital infection acquisition, asymptomatic extragenital screening, and spontaneous resolution of extragenital infections, respectively,

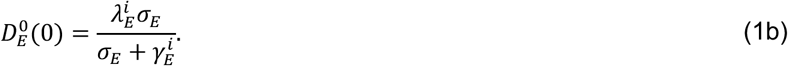

We considered that PrEP initiation may modify these rates of new diagnoses via changes in risk of acquiring each infection (with incidence rate ratio θ*^i^*) and in rates of genital and extragenital screening (with the additive rates *ρ_G_* and ρ*_E_* for PrEP-associated screening):

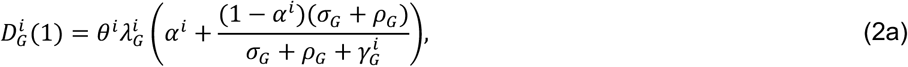

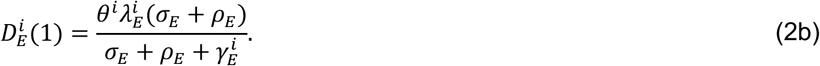

The pathogen-specific IRRs of genital, extragenital, and all diagnoses were thus

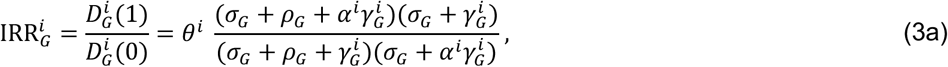

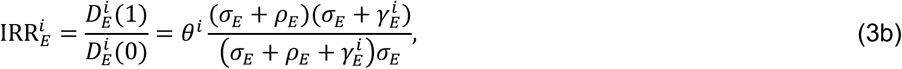

and

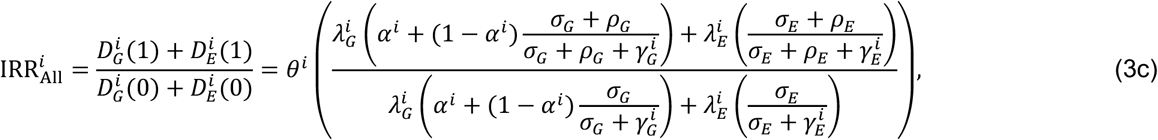

respectively.

In Eq. (3c), identifiability is hindered by product relationships between the unknown parameters θ*^i^* and ρ*_G_*, and θ*^i^* and ρ*_E_*. This prevents joint estimation of the changes in rates of infection and screening associated with PrEP adoption. However, relative changes in rates of diagnoses of genital and extragenital infections provide a formulation independent of θ*^i^* as well as the unobserved parameters 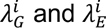:

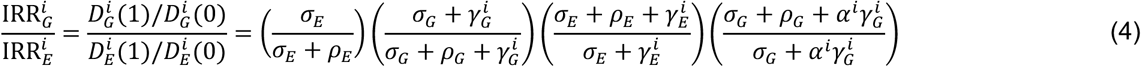

We note that Eq. (4) holds if we extend Eq. (1) and Eq. (2) to consider that providers may have differential likelihood of assigning anatomical site-specific diagnosis codes based on either the site of infection or individuals’ use of PrEP. Defining ω*_k_* as a probability of assigning a site-specific diagnosis code in the scenario of infection at site *k*,

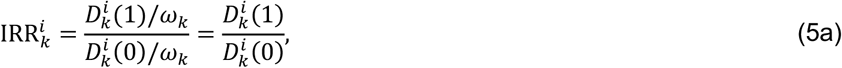

while if we instead consider that ω(*Z*) is the probability of an anatomical site-specific diagnosis given PrEP receipt status

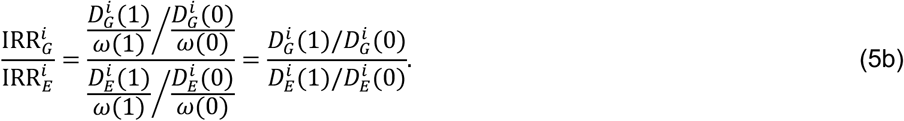

We used our estimates of the ratios 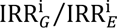 within the study cohort to sample from the distribution of parameters included in Eq. (4), as described below (section below: **Posterior**), defining prior distributions using data from previous studies. Using the fitted values, we compared observations to expectations under the null hypothesis (θ*^i^* = 1), where

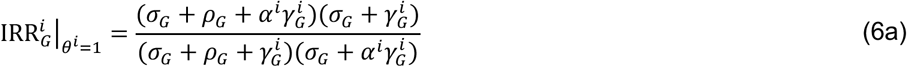

and

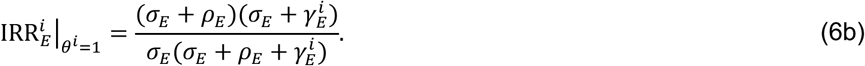

We defined one-sided *p*-values for the hypothesis test that increased rates of diagnoses exceeded expectations based on increases in screening alone as

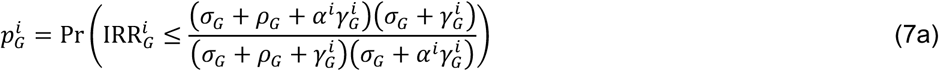

and

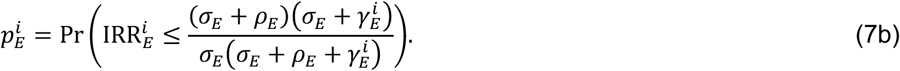

We measured the departure of observed from expected changes in rates of diagnoses, accounting for changes in screening, via

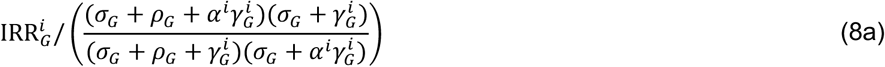

for genital infections, and

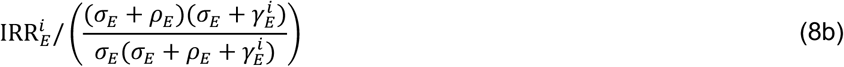

for extragenital infections.

### Posterior

We used Markov Chain Monte Carlo with a Metropolis Hastings step to sample from the joint distribution of the parameters 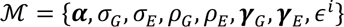. We specified the likelihood ℒ(ℳ|Data) as

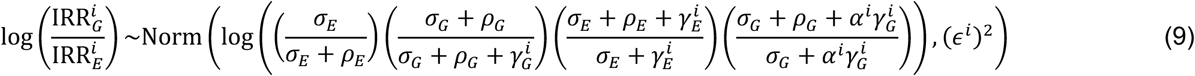

for each pathogen *i*. We defined the posterior as the product of the likelihood contributions for both *N. gonorrhoeae* and *C. trachomatis* as well as the prior probabilities for the individual parameters detailed in the supporting information (**Text S1**). We ran 4 independently-initialized chains for 4 million steps, each, after a burn-in phase of 100,000 steps, thinning chains by a factor of 1/10^th^. We detail further aspects of Markov Chain Monte Carlo sampling in the supporting information (**Text S2**; **Figure S1**).

### Attributable fraction of diagnoses associated with screening

We used our parameter estimates to assess the contribution of changes in screening to estimated changes in incidence of gonorrhea and chlamydia diagnoses after PrEP adoption. Per Eq. (3c) above, 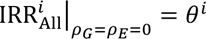, and the attributable fraction of the increase in diagnoses associated with increases in screening was

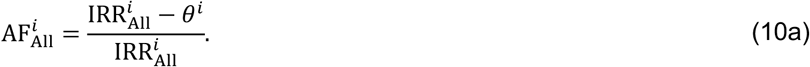

Solving for 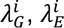, and θ*^i^* via Eqs. (1a), (1b), and (3c), we defined the change in rates of diagnoses associated with screening as 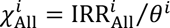, such that

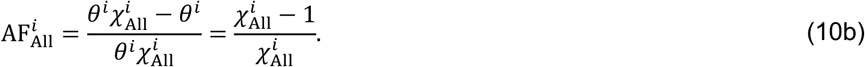

Based on the same framework, we defined 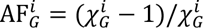 and 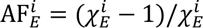.

### Ethics

These analyses of deidentified insurance claims data were considered exempt from review by the University of Alabama at Birmingham Institutional Review Board and the Committee for the Protection of Human Subjects at the University of California, Berkeley.

## RESULTS

### Sample characteristics

Of 11,310 eligible individuals who initiated PrEP during the study period, 9,143 (80.8%) had a second PrEP fill within 365 days of their first and were included in primary analyses (**Table 1**). This cohort contributed a total of 15,377 and 9,826 person-years of follow-up before and after PrEP initiation, respectively (**Table S9**; **Table S10**); mean follow-up time per individual was 1.68 person-years (interquartile range [IQR]: 1.08–2.17) during the control period and 1.07 person-years during the exposure period (IQR: 0.50–1.58). The cohort was geographically and demographically diverse, with the largest proportions aged 30–39 years (*n=*2,613; 28.6%) and residing in the South Atlantic region (*n=*2,370; 25.9%; **Table 1**).

**Table 1:**
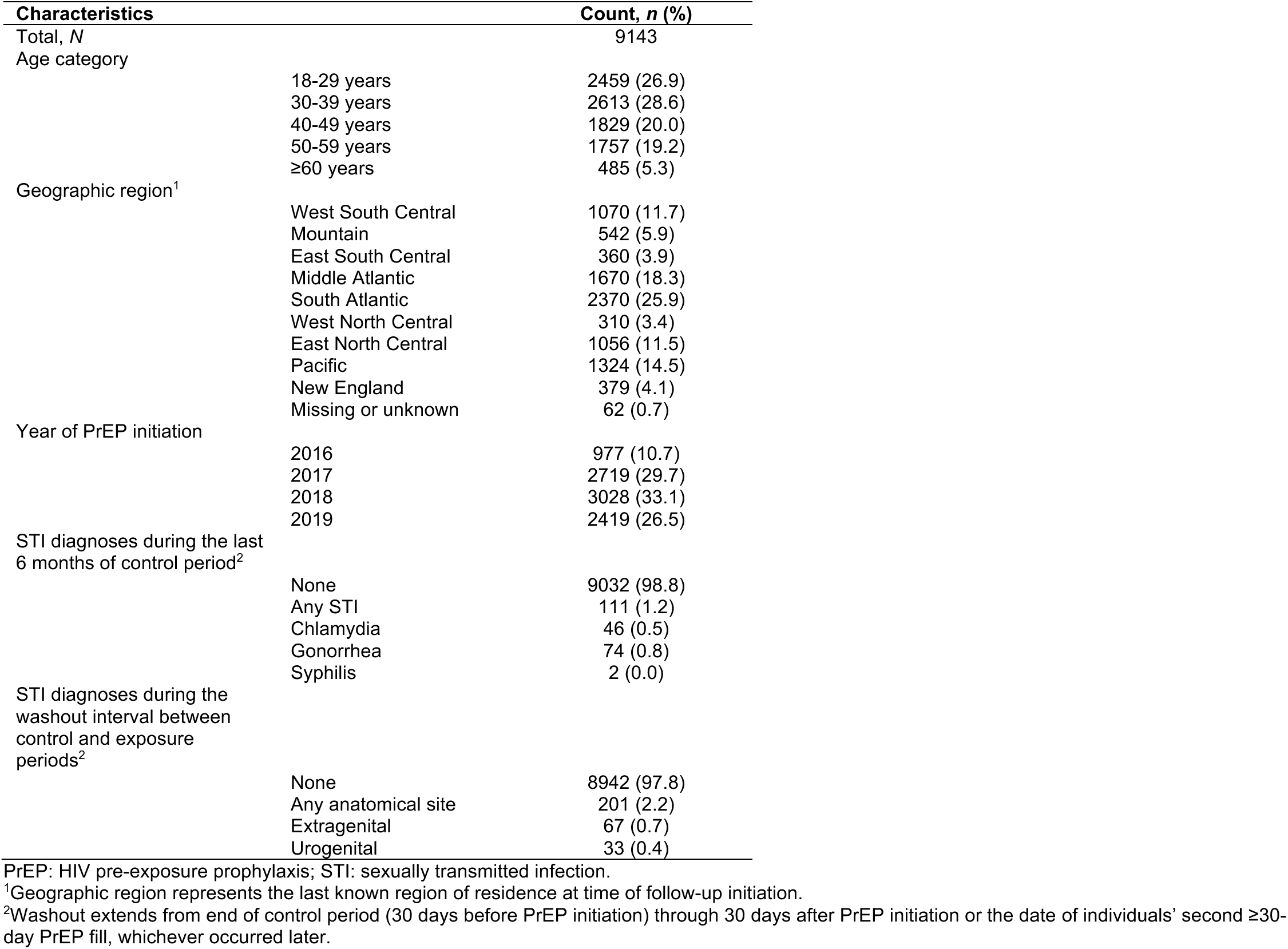
Characteristics of the study population.

| Characteristics |  | Count, *n* (%) |
| --- | --- | --- |
| Total, *N* |  | 9143 |
| Age category | 18-29 years | 2459 (26.9) |
|  | 30-39 years | 2613 (28.6) |
|  | 40-49 years | 1829 (20.0) |
|  | 50-59 years | 1757 (19.2) |
|  | ≥60 years | 485 (5.3) |
| Geographic region<sup>1</sup> | West South Central | 1070 (11.7) |
|  | Mountain | 542 (5.9) |
|  | East South Central | 360 (3.9) |
|  | Middle Atlantic | 1670 (18.3) |
|  | South Atlantic | 2370 (25.9) |
|  | West North Central | 310 (3.4) |
|  | East North Central | 1056 (11.5) |
|  | Pacific | 1324 (14.5) |
|  | New England | 379 (4.1) |
|  | Missing or unknown | 62 (0.7) |
| Year of PrEP initiation | 2016 | 977 (10.7) |
|  | 2017 | 2719 (29.7) |
|  | 2018 | 3028 (33.1) |
|  | 2019 | 2419 (26.5) |
| STI diagnoses during the last 6 months of control period<sup>2</sup> | None | 9032 (98.8) |
|  | Any STI | 111 (1.2) |
|  | Chlamydia | 46 (0.5) |
|  | Gonorrhea | 74 (0.8) |
|  | Syphilis | 2 (0.0) |
| STI diagnoses during the washout interval between control and exposure periods<sup>2</sup> | None | 8942 (97.8) |
|  | Any anatomical site | 201 (2.2) |
|  | Extragenital | 67 (0.7) |
|  | Urogenital | 33 (0.4) |

PrEP: HIV pre-exposure prophylaxis; STI: sexually transmitted infection.
<sup>1</sup>Geographic region represents the last known region of residence at time of follow-up initiation.
<sup>2</sup>Washout extends from end of control period (30 days before PrEP initiation) through 30 days after PrEP initiation or the date of individuals' second ≥30-day PrEP fill, whichever occurred later.

### Rates of STI diagnoses before and after PrEP adoption

The incidence rate of diagnoses with any STI during the control period was 2.14 events per 100 person-years (95% confidence interval [CI]: 1.85–2.42), and 111 (1.2%) individuals received STI diagnoses within the last 6 months of this period (**Table 1**; **Table S11**). Gonorrhea was diagnosed the most frequently (1.32 diagnoses per 100 person-years [95% CI: 1.10–1.53]), followed by chlamydia (0.78 [0.64–0.92]) and syphilis (0.04 [0.01–0.07]). Additionally, 201 individuals (2.2%) received ≥1 STI diagnosis during the washout interval between the end of the control period and PrEP initiation (14.57 [12.40–16.67] diagnoses per 100 person-years), consistent with the expectation that STI diagnoses may be a precipitating factor in the decision to initiate PrEP or may be detected through screening undertaken for PrEP initiation.

Incidence rates of diagnoses with any STI, gonorrhea, chlamydia, and syphilis after PrEP initiation were 8.94 (8.15–9.75), 5.53 (5.00–6.03), 3.38 (2.94–3.85), and 0.03 (0.00–0.07) events per 100 person-years, respectively (**Table S11**). Relative frequencies of urogenital versus extragenital diagnoses of gonorrhea and chlamydia differed in the control and exposure periods. Incidence rates of urogenital STI diagnoses moderately exceeded rates of extragenital diagnoses during individuals’ control period (0.52 [0.41–0.64] versus 0.40 [0.28–0.52] diagnoses per 100 person-years). However, incidence rates after PrEP adoption corresponded to 1.23 (0.99–1.49) urogenital and 2.84 (2.38–3.31) extragenital STI diagnoses per 100 person-years at risk. Rates of urogenital and extragenital diagnoses do not sum to total diagnoses because many infections were diagnosed with anatomically-nonspecific codes. The proportion of diagnoses associated with site-specific ICD-10 code designations did not differ during individuals’ control and exposure periods (41.5% and 43.9%, respectively; two-sided *p*=0.498 via *Z* test).

### Association of PrEP adoption with changes in rates of STI diagnoses

Estimated IRRs for diagnoses with gonorrhea, chlamydia, and either STI after PrEP initiation were 3.47 (2.57–4.72), 3.59 (2.50–5.27), and 3.51 (2.76–4.58), respectively (**Table 2**). We observed similar elevation in rates of diagnoses in analyses subset to individuals with prior STI history (IRR=3.23 [1.18–9.41] for gonorrhea or chlamydia; **Table S12**), and in analyses including individuals who filled only one PrEP prescription (IRR=3.08 [2.44–3.90] for gonorrhea or chlamydia; **Table S13**). Estimated associations were weaker, although directionally consistent, in sensitivity analyses that classified all person-time through PrEP initiation as unexposed person-time (IRR=1.36 [1.08–1.71] for gonorrhea or chlamydia; **Table S14**). We obtained similar results in analyses that limited individuals’ control period to observations within 6 or 12 months before PrEP adoption (IRR=3.10 [2.36–4.07] and 3.44 [2.65–4.51], respectively, for gonorrhea or chlamydia; **Table S15**). Estimated increases in STI risk after PrEP initiation were greater in older age groups in comparison to younger age groups (**Table S16**).

**Table 2:** Association of pre-exposure prophylaxis initiation with diagnosis of sexually transmitted infections.

| Diagnosis |  | Incidence rate ratio (95% confidence interval)<sup>1</sup> |
| --- | --- | --- |
| Gonorrhea or chlamydia | Any site | 3.51 (2.76, 4.58) |
|  | Urogenital | 1.23 (0.71, 2.13) |
|  | Extragenital | 5.98 (3.63, 10.29) |
| Gonorrhea | Any site | 3.47 (2.57, 4.72) |
|  | Urogenital | 1.16 (0.57, 2.43) |
|  | Extragenital | 6.07 (3.43, 11.20) |
| Chlamydia | Any site | 3.59 (2.50, 5.27) |
|  | Urogenital | 1.33 (0.64, 2.96) |
|  | Extragenital | 5.81 (2.97, 12.94) |

<sup>1</sup>Incidence rate ratios were calculated using conditional Poisson regression models, matching on individual, and adjusted for calendar time.

Increases in incidence rates of extragenital STI diagnoses after PrEP initiation exceeded corresponding increases for urogenital infections with the same pathogens. Whereas we estimated 1.16 (0.57–2.43) and 1.33 (0.64–2.96) fold increases in rates of urogenital gonorrhea and chlamydia diagnoses after PrEP initiation, respectively, IRRs for extragenital diagnoses were 6.07 [3.43–11.20] for gonorrhea and 5.81 (2.97–12.94) for chlamydia (**Table 2**). These increases were apparent for both rectal and pharyngeal diagnoses (**Table S17**). The observation of greater IRR estimates for extragenital infections in comparison to urogenital infections held across all sensitivity analyses, and in age-stratified analyses (**Table S18**).

### Changes in rates of STI diagnoses associated with other events

In contrast to observations after PrEP initiation, individuals who received HIV diagnoses experienced 32% (15–46%) lower rates of diagnoses with either gonorrhea or chlamydia thereafter (**Table 3**). Reductions were apparent within the first 6 months after HIV diagnosis (IRR=0.76 [0.60–0.93]) and >6 months after diagnosis (IRR=0.50 [0.36–0.68]). Point estimates suggested greater reductions in incidence of extragenital STI diagnoses in comparison to urogenital diagnoses (IRR=0.43 [0.25–0.75] and IRR=0.78 [0.34–1.57]), respectively) in periods >6 months after HIV infection, and this pattern was apparent for both gonorrhea and chlamydia diagnoses.

**Table 3:** Association of new HIV infection with diagnosis of sexually transmitted infections.

| Diagnosis |  | Incidence rate ratio (95% confidence interval)<sup>1</sup> |  |  |
| --- | --- | --- | --- | --- |
|  |  | *After diagnosis* | *≤6 months after diagnosis* | *>6 months after diagnosis* |
| Gonorrhea or chlamydia | Any site | 0.68 (0.54, 0.85) | 0.76 (0.60, 0.93) | 0.50 (0.36, 0.68) |
|  | Urogenital | 0.85 (0.47, 1.41) | 0.88 (0.49, 1.46) | 0.78 (0.34, 1.57) |
|  | Extragenital | 0.71 (0.47, 1.09) | 0.83 (0.55, 1.22) | 0.43 (0.25, 0.75) |
| Gonorrhea | Any site | 0.73 (0.56, 0.94) | 0.78 (0.61, 1.01) | 0.60 (0.43, 0.85) |
|  | Urogenital | 0.95 (0.43, 1.81) | 0.88 (0.36, 1.66) | 1.10 (0.40, 2.57) |
|  | Extragenital | 0.89 (0.55, 1.43) | 1.00 (0.64, 1.55) | 0.55 (0.28, 1.04) |
| Chlamydia | Any site | 0.61 (0.42, 0.86) | 0.73 (0.52, 0.99) | 0.36 (0.21, 0.56) |
|  | Urogenital | 0.72 (0.31, 1.57) | 0.81 (0.35, 1.64) | 0.51 (0.17, 1.39) |
|  | Extragenital | 0.56 (0.24, 1.20) | 0.68 (0.29, 1.32) | 0.32 (0.11, 0.82) |

<sup>1</sup>Incidence rate ratios were calculated using conditional Poisson regression models, matching on individual, and adjusted for calendar time.

Among individuals meeting the study definition of PrEP discontinuation (≥365 days after the most recent ≥30-day TDF-FTC fill), risk of STI diagnoses during the discontinuation period was comparable to risk during the exposure period (4.13 [2.64–5.97] and 4.16 [1.96–6.92] diagnoses per 100 person-years, respectively; **Table S19**). Compared to these individuals’ rate of 2.02 (1.19–2.92) STI diagnoses per 100 person-years during the control period, incidence rates of diagnoses with either gonorrhea or chlamydia during the exposure period and discontinuation period were 1.91 (0.65–6.34) and 1.92 (0.43–10.25) fold higher, respectively (**Table S20**), suggesting a weaker increase in rates of STI diagnoses than we observed in the full study population.

### Screening-adjusted estimates

Initiating PrEP was associated with, on average, 2.29 (2.18–2.42) additional asymptomatic urogenital STI tests and 1.94 (1.84–2.05) additional extragenital tests annually (**Figure 2**). The expected changes in rates of urogenital versus extragenital infection diagnoses 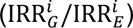 associated only with changes in screening practices were 0.19 (0.07–0.48) for gonorrhea and 0.23 (0.08–0.68) for chlamydia (**Figure 3**). These estimates closely resembled observations in the study cohort: under the null hypothesis of no change in individuals’ risk of acquiring STIs after PrEP initiation, rates of urogenital and extragenital gonorrhea diagnoses were expected to increase 1.08 (1.01–1.25) fold and 5.74 (5.34–6.63) fold, respectively, while for chlamydia, rates of urogenital and extragenital diagnoses were expected to increase 1.20 (1.09–1.37) fold and 5.25 (4.79–6.01) fold, respectively. Observed changes in rates of urogenital gonorrhea and chlamydia diagnoses were 1.08 (0.52–2.27) and 1.13 (0.52–2.50) fold greater than expected under the null hypothesis, respectively (one-sided *p*=0.346 for gonorrhea diagnoses and *p*=0.351 for chlamydia diagnoses). For extragenital infections, observed changes in rates of gonorrhea and chlamydia diagnoses were 1.08 (0.59–2.50) and 1.14 (0.55–2.49) fold greater than expected under the null hypothesis, respectively (one-sided *p*=0.421 and *p=*0.427 for gonorrhea and chlamydia, respectively).

**Figure 2:**
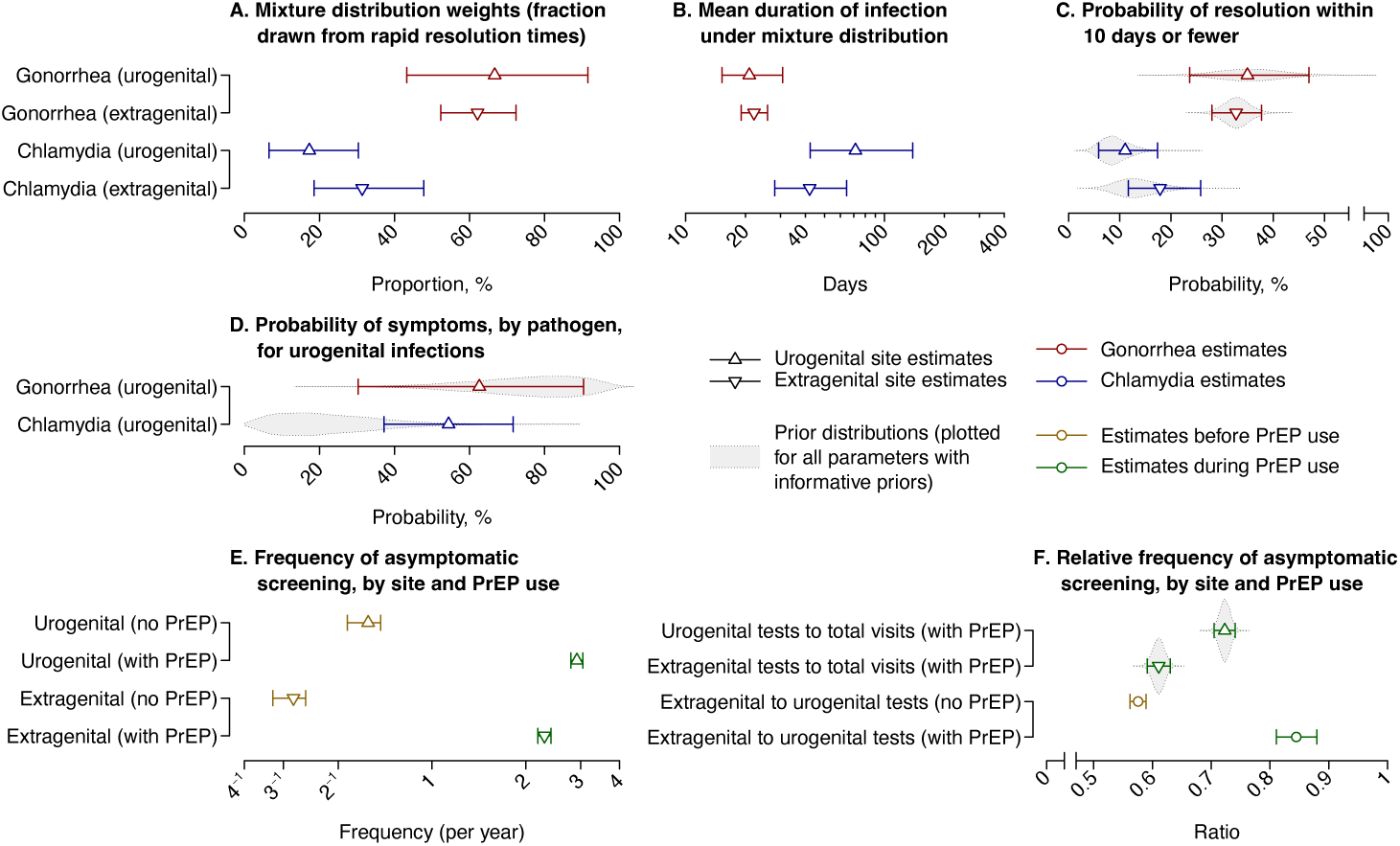
Estimated parameters of gonorrhea and chlamydia natural history and changes in testing behavior associated with initiating HIV pre-exposure prophylaxis (PrEP). Panels illustrate (**A**) distribution weights corresponding to the proportion of incident gonorrhea and chlamydia infections for which durations are drawn from the “short” time to resolution; (**B**) mean times to resolution for untreated gonorrhea and chlamydia infections based on mixture distributions; (**C**) probabilities of resolution within ≤10 days for untreated gonorrhea and chlamydia infections, and (**D**) the proportion of urogenital gonorrhea and chlamydia infections associated with symptoms. Below, we illustrate (**E**) frequencies of asymptomatic urogenital and extragenital screening before and after PrEP initiation, and (**F**) ratios of asymptomatic urogenital and extragenital testing to total PrEP-related healthcare visits, as well as ratios of rates of extragenital to urogenital testing before and after PrEP initiation. For all panels, points indicate mean estimates and lines denote 95% credible intervals. Where priors for parameters were available, we illustrate prior distributions via gray shaded areas.

**Figure 3:**
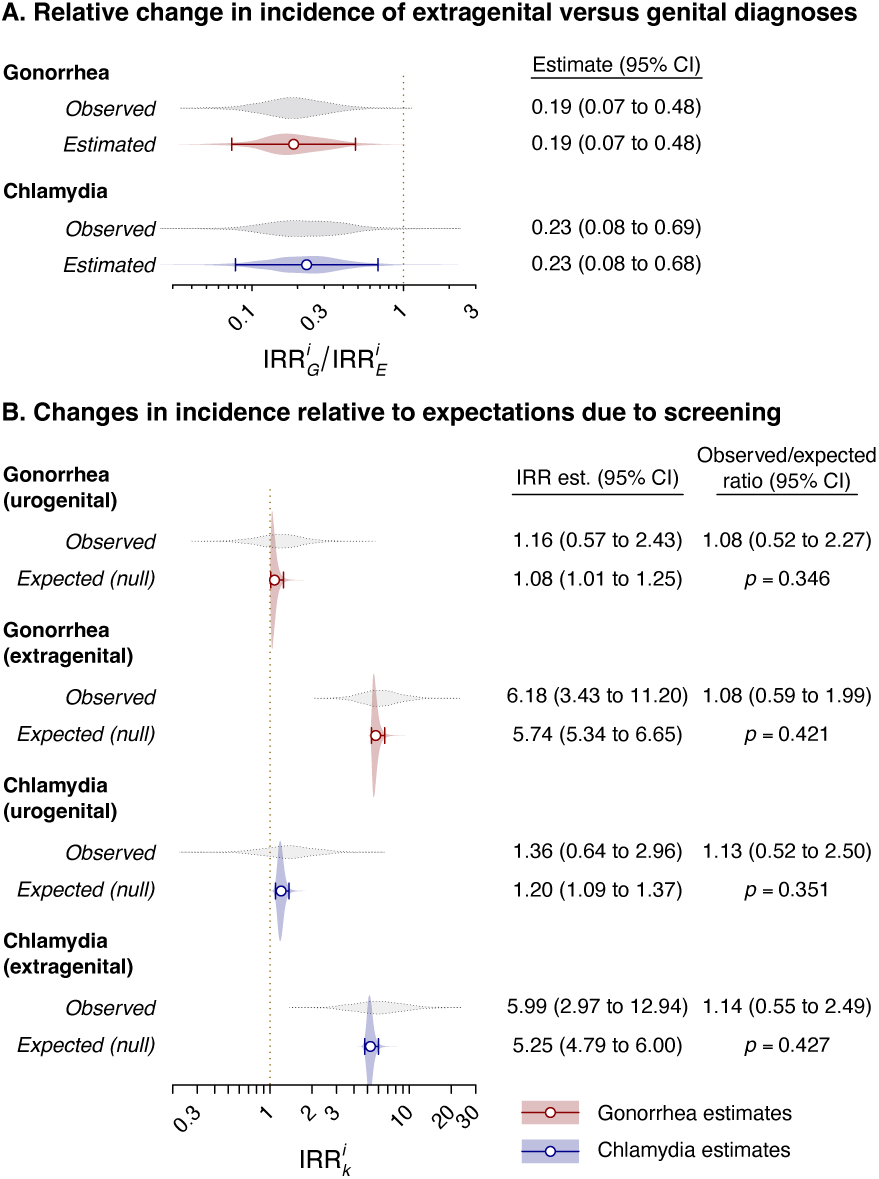
Observations and model-based estimates for changes in gonorrhea and chlamydia incidence after initiating HIV pre-exposure prophylaxis (PrEP). Panels illustrate (**A**) ratios of incidence rate ratios (IRRs) for changes in urogenital relative to extragenital diagnoses, based on both observed data (gray) and estimated model parameters, corresponding to Eq. (4). Panels below (**B**) illustrate observed IRRs of urogenital and extragenital gonorrhea and chlamydia diagnoses associated with PrEP initiation together with expected IRR estimates under the null hypothesis of no change in risk of acquisition, accounting for changes in rates of asymptomatic screening, corresponding to Eq. (6). For all panels, points indicate mean estimates and lines denote 95% credible intervals. Shaded areas illustrate the distribution around estimates. *P*-values listed in panel (**B**) quantify the probability (one-sided) of observing an IRR at least as great as that measured in the study cohort under the null hypothesis, and are computed as specified in Eq. (7).

Increases in asymptomatic screening accounted for 80.7% (75.9–84.6%) of observed increases in gonorrhea diagnoses overall, including 39.2% (–16.4–71.5%) of increases in urogenital diagnoses and 88.9% (82.1–93.9%) of increases in extragenital diagnoses (**Figure 4**). For chlamydia, increases in asymptomatic screening accounted for 80.4% (78.1–83.2%) of observed increases in diagnoses, including 44.8% (–3.6–74.5%%) of increases in urogenital diagnoses and 87.5% (77.0–94.1%) of increases in extragenital diagnoses.

**Figure 4:**
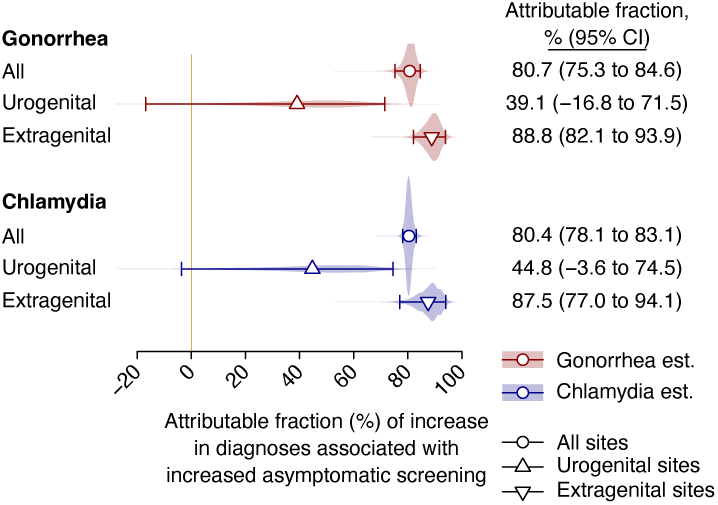
Attributable fraction of the increase in gonorrhea and chlamydia diagnosis risk associated with increases in asymptomatic screening. We plot estimates according to Eq. (10) for infections at all sites and for urogenital and extragenital infections. Shaded areas illustrate the distribution around estimates.

## DISCUSSION

Our findings suggest that higher-frequency asymptomatic STI screening is the primary driver of increased rates of STI diagnoses associated with PrEP initiation in a nationwide cohort of commercially-insured US males. Within our study population, PrEP initiation was associated with >3-fold increases in rates of gonorrhea and chlamydia diagnoses.

However, the increase in rates of diagnoses differed starkly by anatomical site. Whereas rates of extragenital infection diagnoses—mostly detected through screening as infections are typically asymptomatic—increased by similar, >5-fold margins for each infection, urogenital diagnoses more often associated with new-onset symptomatic illness increased by only 16% for gonorrhea and 33% for chlamydia. Accounting for increases in rates of asymptomatic screening, atomical site-specific increases in rates of gonorrhea and chlamydia diagnoses only modestly exceeded expectations under the null hypothesis of no change in infection risk (8-14% greater point estimates than expected). This departure from expected changes in rates of diagnosis did not meet conventional statistical thresholds for rejecting the null hypothesis of no change in individuals’ risk of STIs after PrEP initiation.

Differential increases in rates of STI diagnoses by anatomical site and by pathogen helped to distinguish changes due to screening from changes in infection risk within our study. The association of urogenital *N. gonorrhoeae* infection with greater risk of symptoms than urogenital *C. trachomatis* may explain greater point estimates for increases in urogenital chlamydia diagnoses (IRR=1.33) than urogenital gonorrhea diagnoses (IRR=1.16). In contrast, we observed similar increases in point estimates for extragenital gonorrhea and chlamydia infections (IRR=6.07 and 5.81, respectively), likely reflecting the possibility that changes in asymptomatic screening drive changes in rates of detection. Under distinct analysis frameworks, studies of MSM initiating PrEP in France and Australia have also reported greater increases in rates of chlamydia diagnoses (IRR=1.20–1.78) than gonorrhea diagnoses (IRR=1.05–1.11).^19,20^ Also consistent with our findings, a US study^16^ comparing MSM initiating PrEP to historical controls estimated greater increases in rectal than urogenital diagnoses for both gonorrhea and chlamydia, and greater increases in urogenital chlamydia diagnoses (IRR=2.2) than urogenital gonorrhea diagnoses (IRR=1.5). In a distinct US study reporting test-positive fractions for urethral and extragenital samples tested for *N. gonorrhoeae* and *C. trachomatis*, the proportion of tests associated with positive results did not differ appreciably before and after PrEP initiation.^29^ Our use of a self-matched inference approach and real-world data not limited to patients seen in STI clinic settings augment these prior findings, while our analyses disentangling the role of screening in apparent increases clarifies the likely role of screening in driving similar findings across these studies.

Further supporting the primary role of screening as a driver of changes in rates of STI diagnoses after PrEP adoption, the >3-fold increases in incidence of STI diagnoses observed within our study vastly exceed reported increases in rates of condomless sex among MSM initiating PrEP. Among STI clinic patients in Seattle, PrEP initiation was associated with 10-46% increases in individuals’ likelihood of never using condoms during anal sex, but was not associated with changes in individuals’ total number of male sex partners.^5^ These findings are broadly consistent with data from other studies,^12^ including from cohorts in San Francisco,^17^ Rhode Island,^30^ and non-US settings,^3^ which collectively reported 22-65% increases in participants’ numbers of condomless anal sex partners after PrEP adoption without accompanying changes in total sex partners. The lack of change in total sex partners in these studies further mitigates expected changes in STI risk after PrEP adoption when considering the contribution of oral sex to transmission, especially for *N. gonorrhoeae*.^31,32^ Greater reliance on unprotected anal sex as a transmission pathway for *C. trachomatis* could contribute to higher point estimates for increases in rates of urogenital chlamydia than gonorrhea diagnoses after PrEP initiation in our study and others.^19,20,29^

Similar to PrEP initiation, treatment for newly-diagnosed HIV infection may provide an entry point for engagement with sexual health services including STI testing. However, we identified reductions in individuals’ rate of STI diagnoses after HIV infection diagnoses, contrary to our findings among individuals initiating PrEP. Estimated reductions in STI diagnoses mirror prior evidence that MSM report fewer sex partners and fewer condomless sex acts after new HIV diagnosis.^33–35^ Whereas we identified that increases in rates of STI diagnoses persisted after PrEP discontinuation, it is important to note that individuals who discontinued PrEP experienced modest increases in rates of gonorrhea and chlamydia diagnoses (IRR=1.9) relative to the full study population. This finding may reflect lower adherence to screening within the subgroup of individuals who discontinue PrEP. Lack of new PrEP fills may be a poor indicator of discontinuation if individuals’ TDF-FTC/F-TAF supply extends beyond one year, for instance due to event-driven dosing. Low rates of new sexual partnerships and PrEP consumption may also be a factor in lower rates of screening, and modest increases in STI diagnoses, within the population that appeared to discontinue PrEP.

Our study has several limitations. First, we cannot verify cohort members are MSM based on administrative data alone, although MSM represent the vast majority of PrEP recipients in prior studies and <1% of eligible high-risk heterosexuals use are estimated to use PrEP.^4^ Second, anatomical sites of infection are characterized for only a subset of diagnoses, limiting our statistical power. Third, we sourced prior distributions from distinct study populations, whose patterns of screening and STI natural history may differ. Fourth, we lacked data to distinguish duration of infection at genital and extragenital sites, and did not distinguish between rectal and pharyngeal infections. Last, our analyses captured few syphilis diagnoses, potentially due to our conservative outcome definition requiring a penicillin injection within 10 days of diagnosis.

Findings from our study can inform interpretation of trends in rates of STI diagnoses among MSM over the decade since PrEP implementation. Expanded recommendations for routine extragenital screening among MSM beginning in 2014,^36^ and substantial uptake of PrEP with recommendations for screening of recipients every 3-6 months,^10,14^ may contribute to the >2-fold increases in incidence of gonorrhea diagnoses among US MSM reported between 2012 and 2019.^37^ In view of our results, trends in case-based surveillance of STI incidence should be interpreted cautiously as a basis for informing optimal screening schedules among MSM receiving PrEP. This consideration may have future pertinence if receipt of long-acting (6-month) injectable PrEP is associated with reduced screening frequency among some MSM.

## Data Availability

Raw data from the MarketScan insurance claims databases are available for licensed users. A user license could be obtained by following the instructions at https://marketscan.truvenhealth.com/marketscanportal/.

