## Supporting information for "Enhanced screening and bacterial sexually-transmitted infection diagnoses after HIV pre-exposure prophylaxis initiation"

**Contents of this supplement**

| Item | Title | Page |
|---|---|---|
| Text S1 | Prior distributions. | 3 |
| Text S2 | Markov chain Monte Carlo sampling. | 5 |
| Text S3 | Supplemental references. | 6 |
| Table S1 | National Drug Codes used to identify emtricitabine/tenofovir use. | 7 |
| Table S2 | Codes used to identify HIV infection. | 8 |
| Table S3 | Codes used to identify HIV-associated opportunistic infections. | 9 |
| Table S4 | Codes used to identify Hepatitis B infection. | 10 |
| Table S5 | Codes used to identify needlestick exposure. | 11 |
| Table S6 | Codes used to identify gonococcal infection. | 12 |
| Table S7 | Codes used to identify chlamydia infection. | 13 |
| Table S8 | Codes used to identify syphilis infection. | 14 |
| Table S9 | Observed person-years of follow-up, stratified by analysis population. | 15 |
| Table S10 | Characteristics of populations included in supplemental analyses | 16 |
| Table S11 | Diagnosis rate of sexually transmitted infections before and after PrEP initiation. | 17 |
| Table S12 | Association of PrEP initiation with sexually transmitted infection diagnoses, among individuals with prior sexually transmitted infection diagnoses. | 18 |
| Table S13 | Association of PrEP initiation with sexually transmitted infection diagnoses, including one-time recipients of PrEP. | 19 |
| Table S14 | Association of PrEP initiation with sexually transmitted infection diagnoses, including the enrollment window as unexposed person-time. | 20 |
| Table S15 | Association of PrEP initiation with sexually transmitted infection diagnoses, limiting control periods to 6 and 12 months before PrEP initiation. | 21 |
| Table S16 | Association of PrEP initiation with sexually transmitted infection diagnoses, stratified by age. | 22 |
| Table S17 | Site-specific association of PrEP initiation with extragenital sexually transmitted infection diagnoses. | 23 |
| Table S18 | Ratio of increases in urogenital to extragenital diagnoses. | 24 |
| Table S19 | Incidence rates of sexually transmitted infection diagnoses among individuals discontinuing PrEP during the study period. | 25 |
| Table S20 | Association of PrEP initiation and discontinuation with sexually transmitted infection diagnoses, among individuals discontinuing PrEP during the study period. | 26 |
| Figure S1 | Markov chain Monte Carlo traces. | 27 |

**Text S1: Prior distributions.**

_Symptomatic infection_: To characterize the proportion of urogenital *N. gonorrhoeae* and *C. trachomatis* infections causing symptoms, we defined

$$\alpha^{Ng} \sim \text{Beta}(6,2)$$

$$\alpha^{Ct} \sim \text{Beta}(2,9)$$

using data from a study[1] that employed active surveillance for incident STIs via prospective testing at regularly scheduled intervals, coupled with monitoring of symptomatic disease.

_Urogenital and extragenital screening_: We used observations from ARTnet, a nationwide, survey-based study of US MSM, to characterize screening practices.[2] Data included respondents' reported frequency of sexual health-related visits to care providers after PrEP initiation, $Z^*(1)$, from which we defined

$$Z^*(1) \sim \text{Poisson}(\sigma_G + \rho^*)$$

with $\rho_G = \beta_G \rho^*$, $\rho_E = \beta_E \rho^*$, and $\rho^* \sim \text{Gamma}(10^{-6}, 10^{-6})$. From participants' reported proportion of PrEP-related visits where they received asymptomatic screening tests, $\nu_G$ and $\nu_E$, we defined

$$\beta_G \sim \text{Beta}\left(\sum_j Z_j^* \nu_{G\,j}, \sum_j Z_j^* (1 - \nu_{G\,j})\right)$$

and

$$\beta_E \sim \text{Beta}\left(\sum_j Z_j^* \nu_{E\,j}, \sum_j Z_j^* (1 - \nu_{E\,j}).\right)$$

The same study also provided distributions of the frequency of STI screening over a two-year recall period, among both PrEP recipients and non-recipients. As direct responses on testing frequency prior to PrEP initiation among recipients were not available, we sought to use propensity weighting framework to reconstruct expected rates of prior testing among PrEP recipients from observations among individuals who had not yet initiated PrEP by the time of survey participation. We fit a logistic regression model comparing the following demographic and sexual risk characteristics of PrEP recipients and non-recipients: age group (<18, 18-24, 25-29, 30-34, 35-39, 40-49, ≥50 years), race (White, Black, Asian, American Indian/Alaska Native/Native Hawaiian/Pacific Islander, multiple, or other), Hispanic/non-Hispanic ethnicity, city/metro area, educational attainment (high school, some post-secondary, or post-secondary complete), history of using HIV post-exposure prophylaxis, history of sex with female partners, and number of male sex partners in the preceding year (0, 1-2, 3-5, 6-10, 11-20, 21-40, >40). We used the resulting parameter estimates to generate propensity weights for PrEP receipt among non-recipients, and used these weights to define the contribution of observations from PrEP non-recipients to the prior (multiplying the log prior probability for each observation by the accompanying individual-level weight). As data did not distinguish between genital versus extragenital screening frequencies among individuals not using PrEP, we specified only

$$Z_G(0) \sim \text{Poisson}(\sigma_G).$$

As the ARTnet study did not provide data on individuals' past frequency of STI screening, by site, we used data from a separate study characterizing site-specific STI testing among MSM shortly before PrEP initiation (2010-2012) to inform estimation of $\sigma_E$. The study[6] provided a tabulation of the frequency of genital and extra testing for *C. trachomatis* and *N. gonorrhoeae* among 21,994 MSM across 44,105 visits to STI clinics in 12 US jurisdictions. Assuming (as above) that most or all visits would result in genital testing and a further proportion could result in extragenital testing, we defined the ratio of extragenital screening tests to total genital tests, prior to PrEP initiation, as $\sigma_E / (\lambda_G^i \alpha^i + \sigma_G)$, and defined

$$\frac{\sigma_E}{\lambda_G^i \alpha^i + \sigma_G} \sim \text{Beta}\left(N^i_{\text{Extragenital tests}}, N^i_{\text{Genital tests}} - N^i_{\text{Extragenital tests}}\right)$$

using counts $N$ from the study data. The volume of testing represented in the data vastly exceeded the number of patients enrolled in the ARTnet study ($N_{\text{ARTnet}} = 2{,}515$), rendering MCMC sampling highly sensitive to changes in proposal values for $\sigma_E/\sigma_G$. To assign comparable importance to data from both studies in posterior inference, we down-weighted the prior probability contribution from this study by a factor $N_{\text{ARTnet}}/N^i_{\text{Genital tests}}$ by multiplying this constant by the log prior probability of the term $\sigma_E/(\lambda^i_G\alpha^i + \sigma_G)$. We further defined, as a flat prior,

$$\frac{\sigma_E}{\sigma_G} \sim \text{Beta}(1,1).$$

<u>*Duration of infection*</u>: In addressing the expected duration of infection, we accounted for patterns of *N. gonorrhoeae* and *C. trachomatis* natural history associated with both persistent rapidly-resolving asymptomatic infections. We defined time to spontaneous resolution for pathogen *i* at anatomical site *k* as a mixture distribution,

$$\tau^i_k = \min\left(\tau^i_k(\text{Short}), \tau^i_k(\text{Long})\right)$$

with

$$\tau^i_k(\text{Short}) \sim \text{Exponential}(1/14)$$

and

$$\tau^i_k(\text{Long}) \sim \text{Exponential}(1/365).$$

We defined a free parameter $\pi^i_k \sim \text{Beta}(1,1)$ conveying the proportion of asymptomatic infections expected to result in quickly-resolving infections, so that the probability of infection resolution by day *t* was

$$\Pr\left(\tau^i_k \le t\right) = \pi^i_k(1 - \exp(-t/14)) + \left(1 - \pi^i_k\right)(1 - \exp(-t/965)),$$

and

$$\gamma^i_k = \frac{\pi^i_k}{14} + \frac{1 - \pi^i_k}{365}.$$

To parameterize durations of extragenital infections, we used observations from ExGen,[7] a prospective cohort study of MSM monitoring the duration of untreated extragenital *N. gonorrhoeae* and *C. trachomatis* infections ascertained by retrospective testing of self-collected specimens at weekly intervals. From reported Kaplan-Meier curves, we extracted the number of individuals clearing infection by day *t*=10, $x^i_E(t)$, among all individuals infected at baseline $n^i_E$, and defined

$$x^i_E(t) \sim \text{Binomial}\left(n = n^i_E, p = \Pr\left(\tau^i_E \le t\right)\right).$$

For consistency with real-world circumstances—in which individuals receive diagnoses of gonorrhea or chlamydia based on a single positive nucleic acid amplification test—we used data from the primary ExGen study defining infections as instances with a single positive result. Data used for analyses were limited to anorectal infections, as separate reports on the duration of oropharyngeal infection did not report outcomes based on infections defined from a single positive result. This concern is unlikely to be of importance for *C. trachomatis*, as oropharyngeal infections are uncommon and nearly all extragenital infections observed in the MarketScan cohort are likely to be ascertained from anorectal testing. We note, however, that the ExGen study indicated durations of oropharyngeal gonococcal infection may exceed durations of anorectal infection.[8] Data specified $x^{Ng}_E(t) = 118$, $n^{Ng}_E = 359$, $x^{Ct}_E(t) = 10$, and $n^{Ct}_E = 74$.

For urogenital infections, we summed values of $x^i_G(t)$ and $n^i_G$ across multiple studies which reported these values (separately for gonorrhea[9–11] and chlamydia[9]). Data specified $x^{Ng}_G(t) = 21$, $n^{Ng}_G = 59$, $x^{Ct}_G(t) = 10$, and $n^{Ct}_G = 110$.

Last, we defined $\epsilon^i \sim \text{Gamma}(10^{-6}, 10^{-6})$ for standard error terms in the likelihood formulation (Eq. (9)).

**Text S2: Markov chain Monte Carlo sampling.**

For each of 4 chains, we sampled starting values of parameters according to

$$\sigma^{*} \sim \text{Unif}(0,0.01)$$

$$\beta_G \sim \text{Beta}(1,1)$$

$$\beta_E \sim \text{Beta}(1,1)$$

$$\rho \sim \text{Unif}(0,0.04)$$

$$\frac{\sigma_E}{\sigma_G} \sim \text{Beta}(1,1)$$

$$\pi_G^{Ng} \sim \text{Beta}(1,1)$$

$$\pi_E^{Ng} \sim \text{Beta}(1,1)$$

$$\pi_G^{Ct} \sim \text{Beta}(1,1)$$

$$\pi_E^{Ct} \sim \text{Beta}(1,1)$$

$$\alpha^{Ng} \sim \text{Beta}(1,1)$$

$$\alpha^{Ct} \sim \text{Beta}(1,1)$$

$$\epsilon^{Ng} \sim \text{Unif}(0,5)$$

$$\epsilon^{Ct} \sim \text{Unif}(0,5).$$

For MCMC sampling, we drew from normal proposal distributions with mean equal to 0 and standard deviations equal to 0.001 for $\sigma^{*}$, 0.001 for $\rho$, and 0.01 for all remaining parameters. We proposed updates to 4 parameters per step. We discarded the first 100,000 steps as a burn-in period and sampled from the posterior via 4 million steps thereafter. We saved the state of the chain at every 10$^{th}$ step. We verified the chains sampled from overlapping and stationary distributions via visual inspection (**Figure S1**).

**Text S3: Supplemental references.**

1 Molina J-M, Charreau I, Chidiac C, *et al.* Post-exposure prophylaxis with doxycycline to prevent sexually transmitted infections in men who have sex with men: an open-label randomised substudy of the ANRS IPERGAY trial. *Lancet Infect Dis* 2018; **18**: 308–17.

2 Chandra C, Weiss KM, Kelley CF, Marcus JL, Jenness SM. Gaps in Sexually Transmitted Infection Screening Among Men who Have Sex with Men in Pre-exposure Prophylaxis (PrEP) Care in the United States. *Clin Infect Dis* 2021; **73**: e2261–9.

3 Traeger MW, Cornelisse VJ, Asselin J, *et al.* Association of HIV Preexposure Prophylaxis With Incidence of Sexually Transmitted Infections Among Individuals at High Risk of HIV Infection. *JAMA* 2019; **321**: 1380–90.

4 Nguyen V-K, Greenwald ZR, Trottier H, *et al.* Incidence of sexually transmitted infections before and after preexposure prophylaxis for HIV. *AIDS Lond Engl* 2018; **32**: 523–30.

5 McManus H, Grulich AE, Amin J, *et al.* Comparison of Trends in Rates of Sexually Transmitted Infections Before vs After Initiation of HIV Preexposure Prophylaxis Among Men Who Have Sex With Men. *JAMA Netw Open* 2020; **3**: e2030806.

6 Extragenital Gonorrhea and Chlamydia Testing and Infection Among Men Who Have Sex With Men—STD Surveillance Network, United States, 2010–2012 | Clinical Infectious Diseases | Oxford Academic. https://academic.oup.com/cid/article-abstract/58/11/1564/2895546 (accessed Oct 22, 2025).

7 Barbee LA, Khosropour CM, Soge OO, *et al.* The Natural History of Rectal Gonococcal and Chlamydial Infections: The ExGen Study. *Clin Infect Dis* 2022; **74**: 1549–56.

8 Duration of Pharyngeal Gonorrhea: A Natural History Study | Clinical Infectious Diseases | Oxford Academic. https://academic.oup.com/cid/article/73/4/575/6123922 (accessed Aug 19, 2025).

9 Liere GAFS van, Hoebe CJPA, Dirks JA, Wolffs PF, Dukers-Muijrers NHTM. Spontaneous clearance of urogenital, anorectal and oropharyngeal Chlamydia trachomatis and Neisseria gonorrhoeae in women, MSM and heterosexual men visiting the STI clinic: a prospective cohort study. *Sex Transm Infect* 2019; **95**: 505–10.

10 Teker B, Vries H de, Heijman T, Dam A van, Loeff MS van der, Jongen VW. Spontaneous clearance of asymptomatic anogenital and pharyngeal Neisseria gonorrhoeae: a secondary analysis from the NABOGO trial. *Sex Transm Infect* 2023; **99**: 219–25.

11 Mensforth S, Ayinde OC, Ross J. Spontaneous clearance of genital and extragenital Neisseria gonorrhoeae: data from GToG. *Sex Transm Infect* 2020; **96**: 556–61.

**Table S1: National Drug Codes used to identify emtricitabine/tenofovir use.**

| Code type | Code | Code description |
|---|---|---|
| NDC | 61958-0701-1 | Truvada [emtricitabine/ tenofovir disoproxil fumarate], 200 mg/1, 300 mg/1 |
| NDC | 61958-0703-1 | Truvada [emtricitabine/ tenofovir disoproxil fumarate], 100 mg/1, 150 mg/1 |
| NDC | 61958-0704-1 | Truvada [emtricitabine/ tenofovir disoproxil fumarate], 133 mg/1, 200 mg/1 |
| NDC | 61958-0705-1 | Truvada [emtricitabine/ tenofovir disoproxil fumarate], 167 mg/1, 250 mg/1 |
| NDC | 61958-2005-1 | Descovy [emtricitabine and tenofovir alafenamide] 120 mg/1, 15 mg/1 |
| NDC | 61958-2002-2 | Descovy [emtricitabine and tenofovir alafenamide] 200 mg/1, 25 mg/1 |
| NDC | 61958-2002-1 | Descovy [emtricitabine and tenofovir alafenamide] 200 mg/1, 25 mg/1 |
| NDC | 70771-1620-2 | Emtricitabine / Tenofovir |
| NDC | 70771-1620-3 | Emtricitabine / Tenofovir |
| NDC | 70771-1620-4 | Emtricitabine / Tenofovir |
| NDC | 70771-1620-9 | Emtricitabine / Tenofovir |
| NDC | 70771-1621-2 | Emtricitabine / Tenofovir |
| NDC | 70771-1621-3 | Emtricitabine / Tenofovir |
| NDC | 70771-1621-4 | Emtricitabine / Tenofovir |
| NDC | 70771-1621-9 | Emtricitabine / Tenofovir |
| NDC | 70771-1622-2 | Emtricitabine / Tenofovir |
| NDC | 70771-1622-3 | Emtricitabine / Tenofovir |
| NDC | 70771-1622-4 | Emtricitabine / Tenofovir |
| NDC | 70771-1622-9 | Emtricitabine / Tenofovir |
| NDC | 70771-1709-2 | Emtricitabine / Tenofovir |
| NDC | 70771-1709-3 | Emtricitabine / Tenofovir |
| NDC | 70771-1709-4 | Emtricitabine / Tenofovir |
| NDC | 70771-1709-9 | Emtricitabine / Tenofovir |

**Table S2: Codes used to identify HIV infection.**

| Code type | Code | Code description |
|---|---|---|
| ICD10 | B20.0 | HIV disease resulting in mycobacterial infection |
| ICD10 | B20.1 | HIV disease resulting in other bacterial infections |
| ICD10 | B20.2 | HIV disease resulting in cytomegaloviral disease |
| ICD10 | B20.3 | HIV disease resulting in other viral infections |
| ICD10 | B20.4 | HIV disease resulting in candidiasis |
| ICD10 | B20.5 | HIV disease resulting in other mycoses |
| ICD10 | B20.6 | HIV disease resulting in Pneumocystis jirovecii pneumonia |
| ICD10 | B20.7 | HIV disease resulting in multiple infections |
| ICD10 | B20.8 | HIV disease resulting in other infectious and parasitic diseases |
| ICD10 | B20.9 | HIV disease resulting in unspecified infectious or parasitic disease |
| ICD10 | B21.0 | HIV disease resulting in Kaposi sarcoma |
| ICD10 | B21.1 | HIV disease resulting in Burkitt lymphoma |
| ICD10 | B21.2 | HIV disease resulting in other types of non-Hodgkin lymphoma |
| ICD10 | B21.3 | HIV disease resulting in other malignant neoplasms of lymphoid, haematopoietic and related tissue |
| ICD10 | B21.7 | HIV disease resulting in multiple malignant neoplasms |
| ICD10 | B21.8 | HIV disease resulting in other malignant neoplasms |
| ICD10 | B21.9 | HIV disease resulting in unspecified malignant neoplasm |
| ICD10 | B22.0 | HIV disease resulting in encephalopathy HIV dementia |
| ICD10 | B22.1 | HIV disease resulting in lymphoid interstitial pneumonitis |
| ICD10 | B22.2 | HIV disease resulting in wasting syndrome |
| ICD10 | B22.7 | HIV disease resulting in multiple diseases classified elsewhere |
| ICD10 | B23.0 | Acute HIV infection syndrome |
| ICD10 | B23.1 | HIV disease resulting in (persistent) generalized lymphadenopathy |
| ICD10 | B23.2 | HIV disease resulting in haematological and immunological abnormalities, not elsewhere classified |
| ICD10 | B23.8 | HIV disease resulting in other specified conditions |
| ICD10 | B24 | Unspecified human immunodeficiency virus [HIV] disease |
| ICD10 | O98.7 | HIV complicating pregnancy, childbirth, or the puerperium |
| ICD10 | O98.71X | HIV complicating pregnancy |
| ICD10 | O98.72 | HIV complicating childbirth |
| ICD10 | O98.73 | HIV complicating puerperium |
| ICD10 | Z21 | Asymptomatic HIV infection |

**Table S3: Codes used to identify AIDS-defining opportunistic infections.**

| Code type | Code | Code description |
|---|---|---|
| ICD10 | A07.2 | Cryptosporidiosis |
| ICD10 | A31.0 | Pulmonary mycobacterial infection |
| ICD10 | A31.2 | Disseminated mycobacterium avium |
| ICD10 | B25.X | Cytomegaloviral disease |
| ICD10 | B37.1 | Pulmonary candidiasis |
| ICD10 | B37.81 | Candidal esophagitis |
| ICD10 | B38.X | Coccidioidomycosis |
| ICD10 | B45.X | Cryptococcosis |
| ICD10 | B58.2 | Toxoplasma meningoencephalitis |
| ICD10 | B59 | Pneumocystosis |
| ICD10 | C46.X | Kaposi’s sarcoma |

**Table S4: Codes used to identify Hepatitis B infection.**

| Code type | Code | Code description |
|---|---|---|
| ICD10 | B16x | Acute Hep B |
| ICD10 | B16.0 | Acute hepatitis B with delta-agent with hepatic coma |
| ICD10 | B16.1 | Acute hepatitis B with delta-agent without hepatic coma |
| ICD10 | B16.2 | Acute hepatitis B without delta-agent with hepatic coma |
| ICD10 | B16.9 | Acute hepatitis B without delta-agent and without hepatic coma |
| ICD10 | B17.0 | Acute hepatitis delta infection |
| ICD10 | B17.8 | Other specified acute viral hepatitis |
| ICD10 | B18.0 | Chronic hepatitis B with delta agent |
| ICD10 | B18.1 | Chronic hepatitis B no delta agent |
| ICD10 | B19.1 | Unspecified viral hepatitis B |
| ICD10 | B19.10 | Hepatitis B without hepatic coma |
| ICD10 | B19.11 | Hepatitis B with hepatic coma |
| ICD10 | O98.419 | Viral hepatitis complicating pregnancy, unspecified trimester |
| ICD10 | Z20.5 | Contact with and (suspected) exposure to viral hepatitis |
| ICD10 | Z11.59 | Encounter for screening for other viral diseases |
| ICD10 | Z22.51 | Hepatitis B carrier |

**Table S5: Codes used to identify needlestick exposure.**

| Code type | Code | Code description |
|---|---|---|
| ICD10 | W46 | Contact with hypodermic needle |
| ICD10 | W46.0 | Contact with hypodermic needle |
| ICD10 | W46.1 | Contact with contaminated hypodermic needle |
| ICD10 | W46.0XXA | Contact with hypodermic needle, initial encounter |
| ICD10 | W46.0XXD | Contact with hypodermic needle, subsequent encounter |
| ICD10 | W46.0XXS | Contact with hypodermic needle, sequela |
| ICD10 | W46.1XXA | Contact with contaminated hypodermic needle, initial encounter |
| ICD10 | W46.1XXD | Contact with contaminated hypodermic needle, subsequent encounter |
| ICD10 | W46.1XXS | Contact with contaminated hypodermic needle, sequela |
| ICD10 | Z29.9 | Unspecified prophylaxis |

**Table S6: Codes used to identify gonococcal infection.**

| Code type | Code | Code description | Anatomical site |
|---|---|---|---|
| ICD10 | A54 | Gonococcal infection | Any |
| ICD10 | A54.0 | Gonococcal infection of lower genitourinary tract without periurethral or accessory gland abscess | Urogenital |
| ICD10 | A54.00 | Gonococcal infection of lower genitourinary tract | Urogenital |
| ICD10 | A54.01 | Gonococcal cystitis and urethritis | Urogenital |
| ICD10 | A54.02 | Gonococcal vulvovaginitis | Urogenital |
| ICD10 | A54.03 | Gonococcal cervicitis | Urogenital |
| ICD10 | A54.09 | Other gonococcal infection of lower genitourinary tract | Urogenital |
| ICD10 | A54.1 | Gonococcal infection of lower genitourinary tract with periurethral and accessory gland abscess | Urogenital |
| ICD10 | A54.2 | Gonococcal pelviperitonitis and other gonococcal genitourinary infection | Urogenital |
| ICD10 | A54.21 | Gonococcal infection of kidney and ureter | Urogenital |
| ICD10 | A54.22 | Gonococcal prostatitis | Urogenital |
| ICD10 | A54.23 | Gonococcal infection of other male genital organs | Urogenital |
| ICD10 | A54.24 | Gonococcal female pelvic inflammatory disease | Urogenital |
| ICD10 | A54.29 | Other gonococcal genitourinary infections | Urogenital |
| ICD10 | A54.5 | Gonococcal pharyngitis | Pharyngeal |
| ICD10 | A54.6 | Gonococcal infection of anus and rectum | Rectal |
| ICD10 | A54.9 | Gonococcal infection | Any |

### Table S7: Codes used to identify chlamydia infection.

| Code type | Code | Code description | Anatomical site |
|---|---|---|---|
| ICD10 | A55 | Chlamydial lymphogranuloma | Any |
| ICD10 | A56.00 | Chlamydial infection of lower genitourinary tract | Urogenital |
| ICD10 | A56.01 | Chlamydial cystitis and urethritis | Urogenital |
| ICD10 | A56.02 | Chlamydial vulvovaginitis | Urogenital |
| ICD10 | A56.09 | Other chlamydial infection of lower genitourinary tract | Urogenital |
| ICD10 | A56.11 | Chlamydial female pelvic inflammatory disease | Urogenital |
| ICD10 | A56.19 | Other chlamydial genitourinary infection | Urogenital |
| ICD10 | A56.2 | Chlamydial infection of genitourinary tract, unspecified | Urogenital |
| ICD10 | A56.3 | Chlamydial infection of anus and rectum | Rectal |
| ICD10 | A56.4 | Chlamydial infection of pharynx | Pharyngeal |
| ICD10 | A56.8 | Sexually transmitted chlamydial infection of other sites | Extragenital |
| ICD10 | A74.9 | Chlamydial infection, unspecified | Any |

**Table S8: Codes used to identify syphilis infection.**

| Code type | Code | Code description |
|---|---|---|
| ICD10 | A51 | Early syphilis |
| ICD10 | A51.0 | Primary genital syphilis |
| ICD10 | A51.1 | Primary anal syphilis |
| ICD10 | A51.2 | Primary syphilis of other sites |
| ICD10 | A51.3 | Secondary syphilis of skin and mucous membranes |
| ICD10 | A51.31 | Condyloma latum |
| ICD10 | A51.32 | Syphilitic alopecia |
| ICD10 | A51.39 | Other secondary syphilis of skin |
| ICD10 | A51.4 | Other secondary syphilis |
| ICD10 | A51.41 | Secondary syphilitic meningitis |
| ICD10 | A51.42 | Secondary syphilitic female pelvic disease |
| ICD10 | A51.43 | Secondary syphilitic oculopathy |
| ICD10 | A51.44 | Secondary syphilitic nephritis |
| ICD10 | A51.45 | Secondary syphilitic hepatitis |
| ICD10 | A51.46 | Secondary syphilitic osteopathy |
| ICD10 | A51.49 | Gonococcal infection of eye, unspecified |
| ICD10 | A51.5 | Early syphilis latent |
| ICD10 | A51.9 | Early syphilis unspecified |
| ICD10 | A52 | Late syphilis |
| ICD10 | A52.0 | Cardiovascular and cerebrovascular syphilis |
| ICD10 | A52.00 | Cardiovascular syphilis unspecified |
| ICD10 | A52.01 | Gonococcal infection of musculoskeletal system, unspecified |
| ICD10 | A52.02 | Syphilitic aortitis |
| ICD10 | A52.03 | Syphilitic endocarditis |
| ICD10 | A52.04 | Syphilitic cerebral arteritis |
| ICD10 | A52.05 | Other cerebrovascular syphilis |
| ICD10 | A52.06 | Other syphilitic heart involvement |
| ICD10 | A52.09 | Other cardiovascular syphilis |
| ICD10 | A52.1 | Symptomatic neurosyphilis |
| ICD10 | A52.10 | Symptomatic neurosyphilis, unspecified |
| ICD10 | A52.11 | Tabes dorsalis |
| ICD10 | A52.12 | Other cerebrospinal syphilis |
| ICD10 | A52.13 | Late syphilitic meningitis |
| ICD10 | A52.14 | Late syphilitic encephalitis |
| ICD10 | A52.15 | Late syphilitic neuropathy |
| ICD10 | A52.17 | General paresis |
| ICD10 | A52.19 | Other symptomatic neurosyphilis |
| ICD10 | A52.2 | Asymptomatic neurosyphilis |
| ICD10 | A52.3 | Neurosyphilis, unspecified |
| ICD10 | A52.7 | Other symptomatic late syphilis |
| ICD10 | A52.71 | Late syphilitic oculopathy |
| ICD10 | A52.72 | Syphilis of lung and bronchus |
| ICD10 | A52.73 | Symptomatic late syphilis of other respiratory organs |
| ICD10 | A52.74 | Syphilis of liver and other viscera |
| ICD10 | A52.75 | Syphilis of kidney and ureter |
| ICD10 | A52.76 | Other genitourinary symptomatic late syphilis |
| ICD10 | A52.77 | Syphilis of bone and joint |
| ICD10 | A52.78 | Syphilis of other musculoskeletal tissue |
| ICD10 | A52.79 | Other symptomatic late syphilis |
| ICD10 | A52.8 | Late syphilis latent |
| ICD10 | A52.9 | Late syphilis unspecified |
| ICD10 | A53 | Other and unspecified syphilis |
| ICD10 | A53.0 | Latent syphilis, unspecified as early or late |
| ICD10 | A53.9 | Syphilis, unspecified |
| ICD10 | O98.1 | Syphilis complicating pregnancy, childbirth and the puerperium |
| ICD10 | O98.11 | Syphilis complicating pregnancy |
| ICD10 | O98.111 | Syphilis complicating pregnancy, first trimester |
| ICD10 | O98.112 | Syphilis complicating pregnancy, second trimester |
| ICD10 | O98.113 | Syphilis complicating pregnancy, third trimester |
| ICD10 | O98.119 | Syphilis complicating pregnancy, unspecified trimester |
| ICD10 | O98.12 | Syphilis complicating childbirth |
| ICD10 | O98.13 | Syphilis complicating the puerperium |

**Table S9. Observed person-years of follow-up, stratified by analysis population.**

| Cohort | Observation timeframe | | | |
|---|---|---|---|---|
| | *Control period* | | *Exposure Period* | |
| | Total person-years of follow-up | Average person-years of follow-up per individual in cohort; mean (IQR) | Total person-years of follow-up | Average person-years of follow-up per individual in cohort; mean (IQR) |
| PrEP recipients with ≥2 fills[1] | 15376.58 | 1.68 (1.08, 2.17) | 9825.92 | 1.07 (0.50, 1.58) |
| PrEP recipients with prior STI history[2] | 353.50 | 1.41 (0.92, 1.83) | 255.83 | 1.02 (0.50, 1.42) |
| PrEP recipients, including one-time recipients[3] | 19210.33 | 1.70 (1.08, 2.25) | 11321 | 1.00 (0.42, 1.42) |
| PrEP recipients with discontinuation in use[4] | 1090.42 | 1.36 (0.92, 1.73) | 505.08 | 0.63 (0.25, 0.92) |
| Individuals experiencing new HIV infection during follow-up | 15655.75 | 1.90 (1.25, 2.42) | 10769.25 | 1.31 (0.50, 2.00) |

[1]Primary analysis cohort.
[2]PrEP recipients with ≥1 STI diagnosis within 1 year before PrEP initiation, or during PrEP enrollment window.
[3]All individuals initiating PrEP, including those with only on PrEP fill.
[4]Individuals ≥365 days of follow-up without additional PrEP fills after any ≥30-day fill. Person-time in the exposure period is defined as time after discontinuation of PrEP.

**Table S10: Characteristics of populations included in supplemental analyses.**

| Characteristics | | Count, *n* (%) | | | |
|---|---|---|---|---|---|
| | | *PrEP recipients with prior STI history*[2] | *PrEP recipients with discontinuation in use*[3] | *PrEP recipients, including one-time recipients*[4] | *Individuals experiencing new HIV infection during follow-up* |
| Total, *N* | | 251 | 802 | 11310 | 8246 |
| Age category | | | | | |
| | 18-29 years | 106 (42.2) | 215 (26.8) | 3210 (28.4) | 998 (12.1) |
| | 30-39 years | 73 (29.1) | 238 (29.7) | 3222 (28.5) | 1451 (17.6) |
| | 40-49 years | 36 (14.3) | 136 (17.0) | 2233 (19.7) | 1652 (20.0) |
| | 50-59 years | 30 (12.0) | 175 (21.8) | 2086 (18.4) | 2912 (35.3) |
| | ≥60 years | 6 (2.4) | 38 (4.7) | 559 (4.9) | 1233 (15.0) |
| Geographic region[1] | | | | | |
| | West South Central | 26 (10.4) | 82 (10.2) | 1299 (11.5) | 1068 (13.0) |
| | Mountain | 17 (6.8) | 46 (5.7) | 658 (5.8) | 390 (4.7) |
| | East South Central | 9 (3.6) | 31 (3.9) | 455 (4.0) | 435 (5.3) |
| | Middle Atlantic | 58 (23.1) | 162 (20.2) | 2165 (19.1) | 1178 (14.3) |
| | South Atlantic | 57 (22.7) | 205 (25.6) | 2864 (25.3) | 2974 (36.1) |
| | West North Central | 4 (1.6) | 20 (2.5) | 360 (3.2) | 238 (2.9) |
| | East North Central | 23 (9.2) | 97 (12.1) | 1327 (11.7) | 789 (9.6) |
| | Pacific | 41 (16.3) | 123 (15.3) | 1632 (14.4) | 732 (8.9) |
| | New England | 16 (6.4) | 34 (4.2) | 479 (4.2) | 183 (2.2) |
| | Missing or unknown | 0 (0) | 2 (0.2) | 71 (0.6) | 259 (3.1) |
| Year exposure began | | | | | |
| | 2016 | 7 (2.8) | 149 (18.6) | 1190 (10.5) | 806 (9.8) |
| | 2017 | 90 (35.9) | 397 (49.5) | 3252 (28.8) | 3021 (36.6) |
| | 2018 | 83 (33.1) | 256 (31.9) | 3682 (32.6) | 2102 (25.5) |
| | 2019 | 71 (28.3) | 0 (0) | 3186 (28.2) | 2317 (28.1) |
| STI diagnosed during the last 6 months of control period | | | | | |
| | None | 170 (67.7) | 792 (98.8) | 11177 (98.8) | 7971 (96.7) |
| | Any STI | 81 (32.3) | 10 (1.2) | 133 (1.2) | 275 (3.3) |
| | Chlamydia | 31 (12.4) | 5 (0.6) | 53 (0.5) | 134 (1.6) |
| | Gonorrhea | 58 (23.1) | 5 (0.6) | 89 (0.8) | 166 (2.0) |
| | Syphilis | 2 (0.8) | 0 (0) | 2 (0.0) | 16 (0.2) |
| STI's diagnosed during PrEP enrollment | | | | | |
| | None | 72 (28.7) | 782 (97.5) | 11067 (97.9) | -- |
| | Any anatomical site | 179 (71.3) | 20 (2.5) | 243 (2.1) | -- |
| | Extragenital | 60 (23.9) | 9 (1.1) | 81 (0.7) | -- |
| | Urogenital | 29 (11.6) | 2 (0.2) | 38 (0.3) | -- |

PrEP: HIV pre-exposure prophylaxis; STI: sexually-transmitted infection.
[1]Geographic region represents the last known region of residence before follow-up initiation.
[2]PrEP recipients with ≥1 STI diagnosis within 6 months before PrEP initiation, or during PrEP enrollment window.
[3]Individuals ≥365 days of follow-up without additional PrEP fills after any ≥30-day fill.
[4]All individuals initiating PrEP, including those with only on PrEP fill.

**Table S11: Incidence rate of sexually transmitted infection diagnoses before and after PrEP initiation.**

| Diagnosis | | Events per 100 person-years (95% confidence interval) | | |
|---|---|---|---|---|
| | | *Before initiation* | *End of control period through PrEP initiation* | *After second PrEP fill* |
| Any STI | | | | |
| | Any site | 2.14 (1.85, 2.42) | 14.57 (12.40, 16.67) | 8.94 (8.15, 9.75) |
| Gonorrhea or chlamydia | | | | |
| | Any site | 2.10 (1.82, 2.38) | 14.17 (12.07, 16.27) | 8.91 (8.13, 9.72) |
| | Urogenital | 0.52 (0.41, 0.64) | 2.30 (1.51, 3.08) | 1.23 (0.99, 1.49) |
| | Extragenital | 0.40 (0.28, 0.52) | 4.33 (3.35, 5.38) | 2.84 (2.38, 3.31) |
| Gonorrhea | | | | |
| | Any site | 1.32 (1.10, 1.53) | 9.45 (7.81, 11.09) | 5.53 (5.00, 6.03) |
| | Urogenital | 0.30 (0.21, 0.39) | 1.57 (0.98, 2.17) | 0.72 (0.55, 0.91) |
| | Extragenital | 0.25 (0.16, 0.34) | 3.15 (2.30, 4.07) | 1.86 (1.56, 2.18) |
| Chlamydia | | | | |
| | Any site | 0.78 (0.64, 0.92) | 4.72 (3.67, 5.77) | 3.38 (2.94, 3.85) |
| | Urogenital | 0.22 (0.15, 0.29) | 0.72 (0.33, 1.18) | 0.51 (0.36, 0.68) |
| | Extragenital | 0.16 (0.09, 0.22) | 1.12 (0.66, 1.71) | 0.98 (0.73, 1.27) |
| Syphilis | | | | |
| | Any site | 0.04 (0.01, 0.07) | 0.39 (0.13, 0.72) | 0.03 (0.00, 0.07) |

PrEP: HIV pre-exposure prophylaxis; STI: sexually transmitted infection.
Incidence rates and confidence intervals are computed with an intercept-only generalized linear model with a Poisson link function.

**Table S12: Association of PrEP initiation with sexually transmitted infection diagnoses, among individuals with prior sexually transmitted infection diagnoses.**

| Diagnosis | | Incidence rate ratio (95% confidence interval)[1] |
|---|---|---|
| Gonorrhea or chlamydia | | |
| | Any site | 3.23 (1.18, 9.41) |
| | Urogenital | 2.26 (0.28, 90.07) |
| | Extragenital | 7.02 (1.08, 97.77) |
| Gonorrhea | | |
| | Any site | 3.44 (1.00, 13.57) |
| | Urogenital | 1.85 (0.03, 96.45) |
| | Extragenital | 4.72 (0.53, 247.76) |
| Chlamydia | | |
| | Any site | 2.77 (0.58, 14.61) |
| | Urogenital | – – |
| | Extragenital | – – |

PrEP: HIV pre-exposure prophylaxis. Analyses are subset to PrEP recipients who received ≥1 sexually transmitted infection diagnosis within 6 months before PrEP initiation, or during the PrEP enrollment window.
[1]Incidence rate ratios were calculated using conditional Poisson regression models, matching on individual, and adjusted for calendar month.

**Table S13: Association of PrEP initiation with sexually transmitted infection diagnoses, including one-time recipients of PrEP.**

| Diagnosis | | Incidence rate ratio (95% confidence interval)[1] |
|---|---|---|
| Gonorrhea or chlamydia | | |
| | Any site | 3.08 (2.44, 3.90) |
| | Urogenital | 1.18 (0.69, 1.95) |
| | Extragenital | 4.50 (2.79, 7.16) |
| Gonorrhea | | |
| | Any site | 3.05 (2.30, 4.13) |
| | Urogenital | 1.10 (0.57, 2.08) |
| | Extragenital | 4.75 (2.78, 8.47) |
| Chlamydia | | |
| | Any site | 3.13 (2.20, 4.38) |
| | Urogenital | 1.30 (0.65, 2.61) |
| | Extragenital | 4.11 (2.07, 8.72) |

PrEP: HIV pre-exposure prophylaxis. Analyses include all individuals initiating PrEP, including those with only one PrEP fill.
[1]Incidence rate ratios were calculated using conditional Poisson regression models, matching on individual, and adjusted for calendar month.

**Table S14: Association of PrEP initiation with sexually transmitted infection diagnoses, including time through PrEP initiation as unexposed person-time.**

| Diagnosis | | Incidence rate ratio (95% confidence interval)[1] |
|---|---|---|
| Gonorrhea or chlamydia | | |
| | Any site | 1.36 (1.08, 1.71) |
| | Urogenital | 0.69 (0.41, 1.12) |
| | Extragenital | 1.63 (1.02, 2.51) |
| Gonorrhea | | |
| | Any site | 1.25 (0.94, 1.65) |
| | Urogenital | 0.57 (0.29, 1.05) |
| | Extragenital | 1.40 (0.82, 2.25) |
| Chlamydia | | |
| | Any site | 1.57 (1.14, 2.17) |
| | Urogenital | 0.91 (0.44, 1.85) |
| | Extragenital | 2.27 (1.20, 4.72) |

PrEP: HIV pre-exposure prophylaxis. Analyses include time through PrEP initiation as unexposed person-time.
[1]Incidence rate ratios were calculated using conditional Poisson regression models, matching on individual, and adjusted for calendar month.

**Table S15: Association of PrEP initiation with sexually transmitted infection diagnoses, limiting control periods to 6 and 12 months before PrEP initiation.**

| Diagnosis | | Incidence rate ratio (95% confidence interval) | |
|---|---|---|---|
| | | *Control period 31-180 days before initiation* | *Control period 31-364 days before initiation* |
| Gonorrhea or chlamydia | | | |
| | Any site | 3.10 (2.36, 4.07) | 3.44 (2.65, 4.51) |
| | Urogenital | 1.30 (0.73, 2.32) | 1.30 (0.75, 2.41) |
| | Extragenital | 5.24 (3.14, 9.55) | 5.90 (3.71, 9.95) |
| Gonorrhea | | | |
| | Any site | 3.12 (2.29, 4.31) | 3.33 (2.47, 4.70) |
| | Urogenital | 1.14 (0.51, 2.40) | 1.12 (0.52, 2.31) |
| | Extragenital | 5.82 (3.26, 12.15) | 5.91 (3.36, 10.22) |
| Chlamydia | | | |
| | Any site | 3.06 (2.11, 4.54) | 3.64 (2.52, 5.27) |
| | Urogenital | 1.56 (0.71, 3.60) | 1.59 (0.77, 3.71) |
| | Extragenital | 4.41 (2.05, 10.99) | 5.92 (2.71, 14.37) |

PrEP: HIV pre-exposure prophylaxis.
[1]Incidence rate ratios were calculated using conditional Poisson regression models, matching on individual, and adjusted for calendar month.

**Table S16: Association of PrEP initiation with sexually transmitted infection diagnoses, stratified by age.**

| Diagnosis | | Incidence rate ratio (95% confidence interval)[1] | | |
|---|---|---|---|---|
| | | *<30 years old* | *30-39 years old* | *≥40 years* |
| Gonorrhea or chlamydia | | | | |
| | Any site | 2.79 (1.83, 4.51) | 3.88 (2.58, 6.41) | 4.16 (2.76, 6.11) |
| | Urogenital | 0.94 (0.33, 2.58) | 0.81 (0.26, 3.09) | 2.16 (0.99, 4.74) |
| | Extragenital | 4.53 (2.06, 10.91) | 5.77 (2.62, 15.11) | 8.55 (3.63, 23.05) |
| Gonorrhea | | | | |
| | Any site | 2.70 (1.57, 4.72) | 4.02 (2.53, 7.20) | 4.04 (2.45, 6.87) |
| | Urogenital | 0.63 (0.15, 2.32) | 1.05 (0.25, 5.13) | 2.78 (0.91, 8.06) |
| | Extragenital | 4.77 (2.02, 14.08) | 5.19 (1.82, 15.72) | 10.90 (3.79, 39.76) |
| Chlamydia | | | | |
| | Any site | 2.96 (1.44, 5.98) | 3.65 (1.92, 7.13) | 4.34 (2.52, 8.20) |
| | Urogenital | 2.11 (0.37, 13.16) | 0.54 (0.07, 3.44) | 1.71 (0.51, 5.45) |
| | Extragenital | 4.04 (1.08, 17.70) | 7.83 (1.82, 70.49) | 6.51 (2.03, 28.41) |

PrEP: HIV pre-exposure prophylaxis. Eligibility for each analysis is defined by individuals' age on December 31, 2019.
[1]Incidence rate ratios were calculated using conditional Poisson regression models, matching on individual, and adjusted for calendar month.

**Table S17: Site-specific association of PrEP initiation with extragenital sexually transmitted infection diagnoses.**

| Diagnosis | | Incidence rate ratio (95% confidence interval)[1] |
|---|---|---|
| Gonorrhea or chlamydia | | |
| | Rectal | 5.94 (3.19, 12.47) |
| | Pharyngeal | 6.67 (3.14, 14.85) |
| Gonorrhea | | |
| | Rectal | 6.62 (3.14, 17.97) |
| | Pharyngeal | 6.18 (2.90, 13.53) |
| Chlamydia | | |
| | Rectal | 5.30 (2.24, 14.67) |
| | Pharyngeal | 18.65 (0.90, ∞) |

[1]Incidence rate ratios were calculated using conditional Poisson regression models, matching on individual, and adjusted for calendar month.

**Table S18: Ratio of increases in urogenital to extragenital diagnoses.**

| Diagnosis | Outcome | Ratio of IRR estimates, urogenital to extragenital (95% CI)[1] |
|---|---|---|
| PrEP recipients with ≥2 fills[2] | | |
| | Gonorrhea or chlamydia | 0.21 (0.09, 0.42) |
| | Gonorrhea | 0.19 (0.07, 0.48) |
| | Chlamydia | 0.23 (0.08, 0.69) |
| PrEP recipients with prior STI history[3] | | |
| | Gonorrhea or chlamydia | 0.32 (0.01, 20.46) |
| | Gonorrhea | 0.39 (0.00, 28.45) |
| | Chlamydia | – – |
| PrEP recipients, including one-time recipients[4] | | |
| | Gonorrhea or chlamydia | 0.26 (0.13, 0.54) |
| | Gonorrhea | 0.23 (0.10, 0.56) |
| | Chlamydia | 0.32 (0.12, 0.83) |
| Analyses including time through PrEP initiation as unexposed person-time[5] | | |
| | Gonorrhea or chlamydia | 0.42 (0.21, 0.82) |
| | Gonorrhea | 0.41 (0.18, 0.90) |
| | Chlamydia | 0.40 (0.14, 1.02) |
| Analyses limiting control period to 31-180 days before PrEP initiation | | |
| | Gonorrhea or chlamydia | 0.25 (0.10, 0.53) |
| | Gonorrhea | 0.20 (0.07, 0.50) |
| | Chlamydia | 0.35 (0.11, 1.15) |
| Analyses limiting control period to 31-364 days before PrEP initiation | | |
| | Gonorrhea or chlamydia | 0.22 (0.10, 0.48) |
| | Gonorrhea | 0.19 (0.07, 0.49) |
| | Chlamydia | 0.27 (0.09, 0.82) |

IRR: Incidence rate ratio. PrEP: HIV pre-exposure prophylaxis; STI: sexually transmitted infection.
[1]Incidence rate ratios were calculated using conditional Poisson regression models, matching on individual, and adjusted for calendar month. Values listed in the table are the ratio of the incidence rate ratio for urogenital diagnoses to the incidence rate ratio for extragenital diagnoses.
[2]Primary analysis cohort.
[3]PrEP recipients with ≥1 STI diagnosis within 1 year before PrEP initiation, or during PrEP enrollment window.
[4]All individuals initiating PrEP, including those with only on PrEP fill.
[5]Analyses include time through PrEP initiation as unexposed person-time.

**Table S19: Incidence rates of sexually transmitted infection diagnoses among individuals discontinuing PrEP during the study period.**

| Diagnosis | | Events per 100 person-years (95% confidence interval) | | |
|---|---|---|---|---|
| | | *Control period* | *PrEP period* | *Discontinuation period* |
| Any STI | | | | |
| | Any site | 2.02 (1.19, 2.92) | 4.13 (2.64, 5.97) | 4.16 (1.96, 6.92) |
| Gonorrhea or chlamydia | | | | |
| | Any site | 1.93 (1.10, 2.83) | 4.13 (2.64, 5.97) | 4.16 (1.96, 6.92) |
| | Urogenital | 0.55 (0.18, 1.00) | 0.66 (0.19, 1.22) | 0.79 (0.00, 1.95) |
| | Extragenital | 0.00 (0.00, 0.00) | 0.85 (0.28, 1.43) | 1.19 (0.19, 2.57) |
| Gonorrhea | | | | |
| | Any site | 1.10 (0.55, 1.73) | 2.91 (1.75, 4.19) | 2.57 (1.16, 4.34) |
| | Urogenital | 0.28 (0.00, 0.57) | 0.56 (0.19, 1.02) | 0.20 (0.00, 0.61) |
| | Extragenital | 0.00 (0.00, 0.00) | 0.66 (0.19, 1.22) | 0.99 (0.19, 2.10) |
| Chlamydia | | | | |
| | Any site | 0.83 (0.36, 1.41) | 1.22 (0.56, 2.00) | 1.58 (0.59, 2.87) |
| | Urogenital | 0.28 (0.00, 0.65) | 0.09 (0.00, 0.29) | 0.59 (0.00, 1.40) |
| | Extragenital | 0.00 (0.00, 0.00) | 0.19 (0.00, 0.47) | 0.20 (0.00, 0.78) |
| Syphilis | | | | |
| | Any site | 0.09 (0.00, 0.28) | 0.00 (0.00, 0.00) | 0.00 (0.00, 0.00) |

Incidence rates and confidence intervals are computed with an intercept-only generalized linear model with a Poisson link function.
The discontinuation period is defined as beginning 365 days after individuals' last fill of a ≥30-day supply of TDF-FTC/F-TAF, and ending at any new TDF-FTC/F-TAF fill; diagnosis with hepatitis B virus infection, HIV infection, or AIDS-defining illness; death; disenrollment (without re-enrollment within <60 days); or December 31, 2019.

**Table S20: Association of PrEP initiation and discontinuation with sexually transmitted infection diagnoses, among individuals discontinuing PrEP during the study period.**

| Diagnosis | | Incidence rate ratio (95% confidence interval)[1] | | |
|---|---|---|---|---|
| | | *PrEP period, relative to control period* | *Discontinuation period[2], relative to control period* | *Discontinuation period[2], relative to PrEP period* |
| Gonorrhea or chlamydia | | | | |
| | Any site | 1.91 (0.65, 6.34) | 1.92 (0.43, 10.25) | 1.01 (0.46, 2.22) |
| | Urogenital | 1.68 (0.16, 22.74) | 3.30 (0.00, 106.11) | 1.96 (0.00, 11.87) |
| | Extragenital | – – | – – | 1.18 (0.07, 5.44) |
| Gonorrhea | | | | |
| | Any site | 2.85 (0.86, 11.60) | 3.17 (0.67, 18.68) | 1.11 (0.49, 2.37) |
| | Urogenital | 5.06 (0.30, ∞) | 4.99 (0.00, ∞) | 0.99 (0.00, 6.64) |
| | Extragenital | – – | – – | 1.64 (0.17, 9.32) |
| Chlamydia | | | | |
| | Any site | 0.89 (0.16, 5.78) | 0.75 (0.08, 9.89) | 0.84 (0.24, 3.05) |
| | Urogenital | 0.17 (0.00, 10.69) | 0.84 (0.00, ∞) | 4.95 (0.00, ∞) |
| | Extragenital | – – | – – | 0.35 (0.00, 7.86) |

[1]Incidence rate ratios were calculated using conditional Poisson regression models, matching on individual, and adjusted for calendar month.
[2]Discontinuation periods begin 365 days after the most recent ≥30-day PrEP prescription fill and end at any new fill, death, disenrollment, end of study period, or diagnosis with hepatitis B virus infection, HIV infection, or AIDS-defining illness.

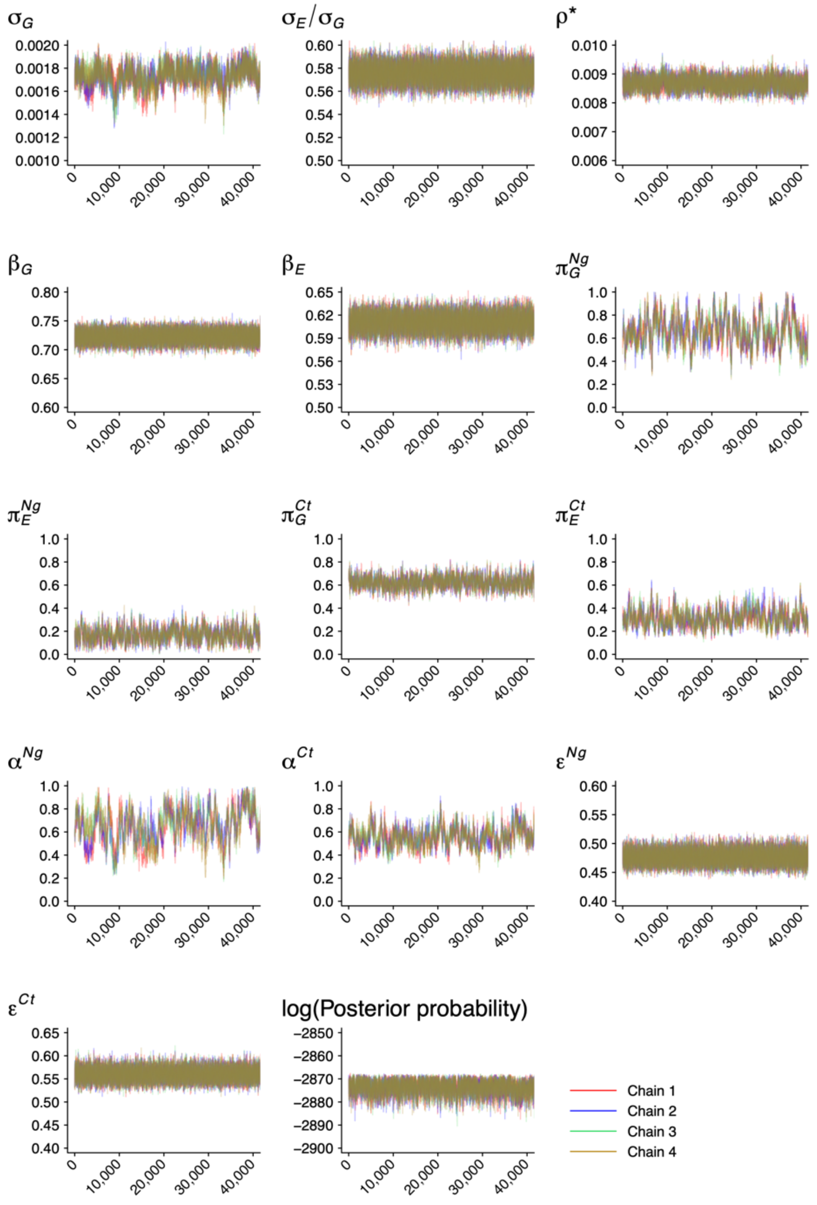

**Figure S1: Markov chain Monte Carlo trace plots.** We illustrate parameter draws across 4,000,000 steps (thinned by a factor of 100) for each of 4 independently-initialized chains after a burn-in of 100,000 iterations.
